# Transmission of SARS-CoV-2 by children and young people in households and schools: a meta-analysis of population-based and contact-tracing studies

**DOI:** 10.1101/2021.12.14.21267713

**Authors:** Russell Viner, Claire Waddington, Oliver Mytton, Robert Booy, Joana Cruz, Joseph Ward, Shamez Ladhani, Jasmina Panovska-Griffiths, Chris Bonell, G.J. Melendez-Torres

## Abstract

**Background:** The role of children and young people (CYP) in transmission of SARS-CoV-2 in household and educational settings remains unclear. We undertook a systematic review and meta-analysis of contact-tracing and population-based studies at low risk of bias.

**Methods:** We searched 4 electronic databases on 28 July 2021 for contact-tracing studies and population-based studies informative about transmission of SARS-CoV-2 from 0-19 year olds in household or educational settings. We excluded studies at high risk of bias, including from under-ascertainment of asymptomatic infections. We undertook multilevel random effects meta-analyses of secondary attack rates (SAR: contact-tracing studies) and school infection prevalence, and used meta-regression to examine the impact of community SARS-CoV-2 incidence on school infection prevalence.

**Findings:** 4529 abstracts were reviewed, resulting in 37 included studies (16 contact-tracing; 19 population studies; 2 mixed studies). The pooled relative transmissibility of CYP compared with adults was 0.92 (0.68, 1.26) in adjusted household studies. The pooled SAR from CYP was lower (p=0.002) in school studies 0.7% (0.2, 2.7) than household studies (7.6% (3.6, 15.9). There was no difference in SAR from CYP to child or adult contacts. School population studies showed some evidence of clustering in classes within schools. School infection prevalence was associated with contemporary community 14-day incidence (OR 1.003 (1.001, 1.004), p<0.001).

**Interpretation:** We found no difference in transmission of SARS-CoV-2 from CYP compared with adults within household settings. SAR were markedly lower in school compared with household settings, suggesting that household transmission is more important than school transmission in this pandemic. School infection prevalence was associated with community infection incidence, supporting hypotheses that school infections broadly reflect community infections. These findings are important for guiding policy decisions on shielding, vaccination school and operations during the pandemic.

**Funding:** No funding obtained.

## Background

The role of children and young people (CYP) in transmission of SARS-CoV-2 remains unclear, in both households and child-specific settings, such as schools and nurseries.[1] Observations of low incidence of symptomatic infection in CYP early in the pandemic led to assumptions that they played a very limited role in infection or transmission. This view has been challenged by the recognition that high proportions of asymptomatic infections in CYP led to low ascertainment of infections in this age-group,[1] particularly when testing capacity was limited. Findings from some large contact-tracing studies (contact-tracing studies)[2] have suggested CYP do play an important role in household transmission. In educational settings, whilst outbreaks have been reported in day-care nurseries,[3] schools[4-6] and school-like residential camps,[7, 8] a number of population-based school studies have found evidence of limited transmission especially between children.[9, 10] It remains unclear the extent to which cases and outbreaks in schools reflect transmission in schools or the wider community.

Epidemiological studies that can provide useful information about transmission with the lowest risk of bias include contact-tracing studies with active follow-up and testing of all contacts regardless of symptoms and population-based studies which test all members of the population regardless of symptoms. Population-based studies are informative about prevalence across age-groups and risk factors for infection, and may provide information about clustering or timing of infection within a setting (e.g. households or schools). Studies have shown that children under 10-12 years have lower susceptibility to SARS-CoV-2 infection than adults, although the risk in teenagers appears to be closer to young adults.[11] However CYP also tend to have the highest social mixing rates across society, including during the pandemic,[12] and transmission is a complex interaction of viral properties, susceptibility, social mixing and population age structures. For these reasons, studies of incidence of symptomatic infection in CYP provide a weak basis for inference around children’s role in transmission. [11]

Over 18 months into the COVID-19 pandemic, there are only now sufficient data to allow meta-analysis of relevant data only including studies at low risk of bias. Existing systematic reviews are now outdated, including only data from early in the pandemic,[13-18] and are critically biased by their inclusion of studies which systematically under-ascertained asymptomatic infections in CYP. A large literature has since been published, including several population-based studies of CYP within schools.[9, 10] Many of these date from late 2020 or early 2021 when schools had extensive mitigation measures in place that are hypothesized to reduce transmission within schools, as does reducing attendance during periods of hybrid in-person and online learning, yet data on the effects of such measures are lacking.[19, 20]

We undertook a systematic review and meta-analysis of high quality epidemiological studies published during the first 18 months of the pandemic (Jan 2020-July 2021) to answer the following questions: (a) To what extent do CYP under 20 years of age transmit SARS-CoV-2 to other CYP and to adults in household and child-specific (e.g. educational) settings?; (b) how does transmission differ between household and educational settings?; and (c) is community infection incidence associated with prevalence of or transmission of infection within educational settings?

## Methods

The search was undertaken using a protocol registered with Prospero registry (CRD42021222276).

### Search strategy

We searched four electronic databases (PubMed; medRxiv; COVID-19 Living Evidence database; Europe PMC) to 28 July 2021. The search terms for PubMed were (“COVID-19”[Text Word] OR “2019-nCoV”[Text Word] OR “SARS-CoV-2”[Text Word]) AND (“child*”[All Fields] OR “infant*”[All Fields]) AND (“disease transmission, infectious”[MeSH Terms] OR “epidemiology”[MeSH Terms] OR “schools”[MeSH Terms]) with terms for other databases shown in Appendix Table 1.

We defined children and young people as being < 20 years of age, but note that different studies used different age-ranges across childhood. We did not limit studies by date or language. The reference lists of identified relevant reviews were checked for additional likely studies. Studies were also identified through other systematic reviews and the professional networks of the authors.

### Eligibility

We searched for contact-tracing studies and community incidence studies to answer questions a) and b), and school incidence or prevalence studies to answer question c). We included published or unpublished reports of studies of SARS-CoV-2 infection of the following types:

a. Contact-tracing studies informative about transmission from primary or index cases aged 0-19 years separately to adult index cases and which identified and tested all contacts regardless of symptoms
b. Population-based studies that were either:
  i. longitudinal incidence studies in any setting which reported or modelled transmission chains between 0-19 year olds and others
  ii. studies of prevalence or incidence in 0-19 year olds in child-specific settings (e.g. day-care, nurseries or schools) using either longitudinal or cross-sectional designs

We only included studies which identified SARS-CoV-2 infection through RT-PCR on oral or nasal samples or through established serological methods. We did not include studies which used less well validated methods such as rapid antigen tests, stool samples[21] or wastewater methods.

We excluded studies of transmission from single individuals or within single institutions; modelling studies that did not provide observational data; studies of vertical transmission; systematic reviews; studies only of school staff; and biological studies of transmission dynamics such as viral load, viral shedding or aerosolization. We excluded ecological level studies of the impact of school opening or closing on community transmission as this has been examined in a separate review.[22]

We excluded studies judged to be at critical risk of bias relating to inadequate ascertainment of asymptomatic infections in CYP. We, therefore, excluded:

1. contact-tracing studies which only tested symptomatic contacts, tested low proportions of recruited contacts or provided insufficient information to judge completeness of contact testing.
2. population studies where infection was identified only by testing of symptomatic individuals or recruitment from clinical settings
3. non-representative population studies due to limited sampling of the target population e.g. where testing was only performed in low proportions of participants

### Study selection

Titles and abstracts of identified studies were reviewed for potential eligibility by one researcher (RV). Those potentially eligible were retrieved in full-text and reviewed independently by 2 researchers (RV and CW or OM) for eligibility and quality.

### Outcomes and data extraction

Outcomes of interest were:

1. From contact-tracing studies: secondary attack rates (SAR) by age of index cases (<18-20 years compared adults) in contact-tracing studies. SAR by age of contact, SAR from adult index cases and effect estimates for adjusted transmission models from CYP were also extracted where data allowed.
2. From population-based studies:
  a. School studies: prevalence or seroprevalence of SARS-CoV-2 infection and presence of clustering (frequency of occurrence of >2 cases) of infection within settings. We also extracted data on school attendance (see below under meta-regression)
  b. longitudinal incidence studies: effect estimates for transmission models from CYP aged 0-19 years.

Data from each study were extracted to a spreadsheet and checked for accuracy by four reviewers (RV, JC, CW and JW). Source of data in each study are shown in Appendix Table 2. We approached authors for further data where necessary.

### Quality and bias evaluation

Methodological quality was independently assessed by two authors (RV and CW) using a score adapted from previously published quality assessment tools[23-26] for prevalence, cohort and case-control studies (see Appendix for details and Appendix Tables 3 and 4). Only studies of high and medium quality at low risk of bias were included in these analyses.

### Data synthesis and analysis

Studies were included in random effects meta-analyses and meta-regressions using a multilevel framework. This accounted for many studies collecting multiple rounds of data collection over time or for studies providing data for CYP age-groups (e.g. primary or secondary students). Analyses used the *metafor* package in R, using log-transformed proportions.

For contact-tracing studies, meta-analyses were undertaken of secondary attack rate (SAR) from index children grouped by setting, age of index child and age of contact. Meta-analysis comparing SAR from child index cases with SAR from adult index cases was undertaken first using raw SAR data and then using estimates of relative transmissibility from adjusted transmission models where data were provided.

For school population-based studies, we first undertook separate meta-analyses of studies providing prevalence and seroprevalence data grouped by age-group. We then used meta-regression to examine associations of school prevalence with:

1. Community 14-day incidence of SARS-CoV-2 across the study period and for the one and two months prior (see Appendix Table 5 for data and sources)
2. School attendance (% face-to-face) in each study (Appendix Table 6). Attendance was measured at the measurement-round level as this varied within a study over time.

We also undertook a post-hoc analysis to examine whether the use of nasopharyngeal or oral swab compared with saliva or gargle sample influenced estimates.

### Role of the funding source

No funding obtained for these analyses.

### Ethics

Ethics permission not required for these secondary analyses of published data.

## Results

The PRISMA flow diagram is shown in Figure 1. Titles and abstracts of 4511 articles were reviewed from electronic databases. Two additional studies were identified through searching citation lists and 16 through professional networks. 336 were assessed in full-text and 89 articles were judged potentially eligible. 45 studies (46 articles) were excluded as being at critical risk of bias (see Appendix Table 7). Characteristics of the 37 included studies (described in 43 articles, some of which describe later rounds of a study) are shown in Table 1.

**Figure 1:**
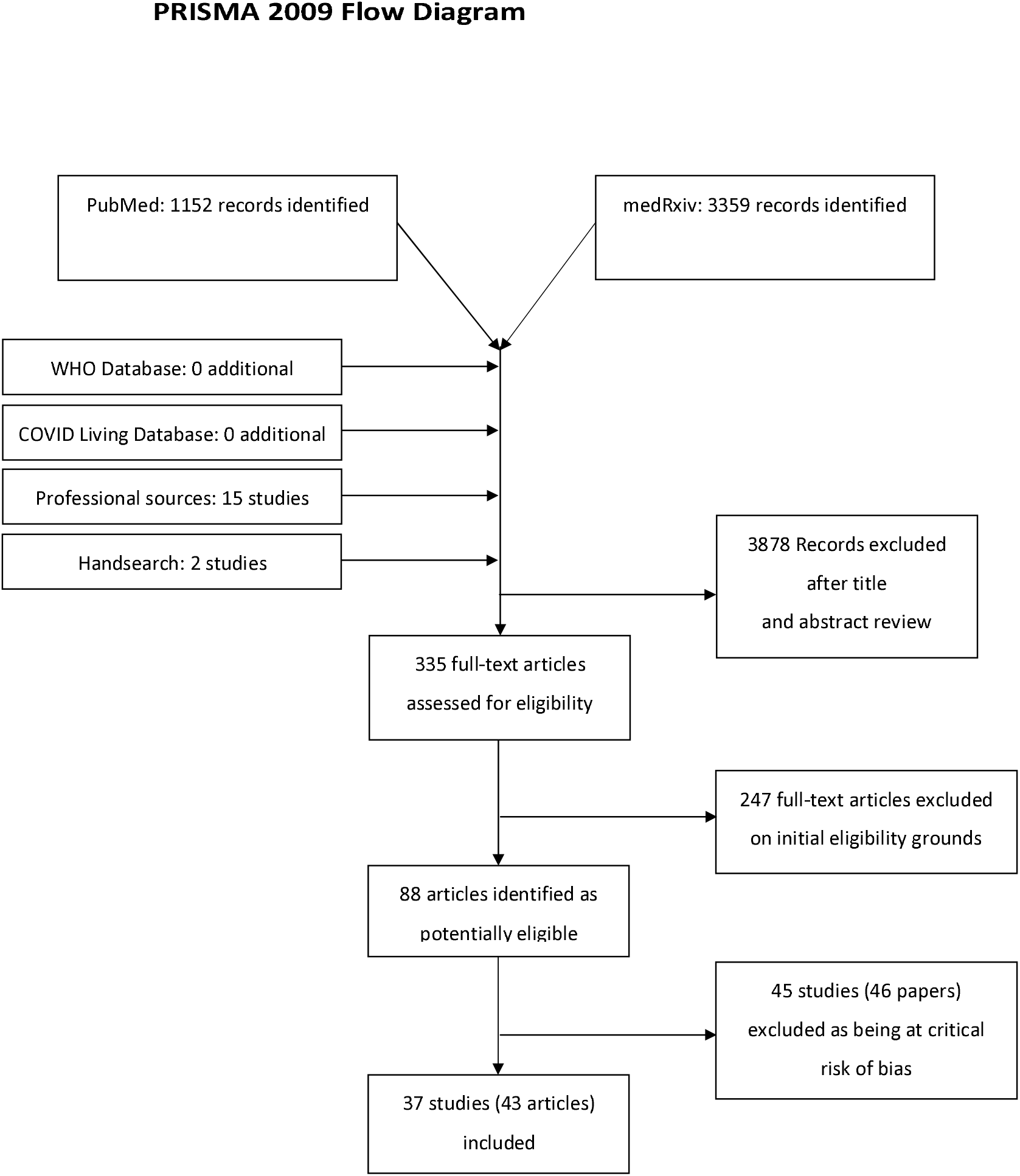
FLOW diagram

**Table 1:**
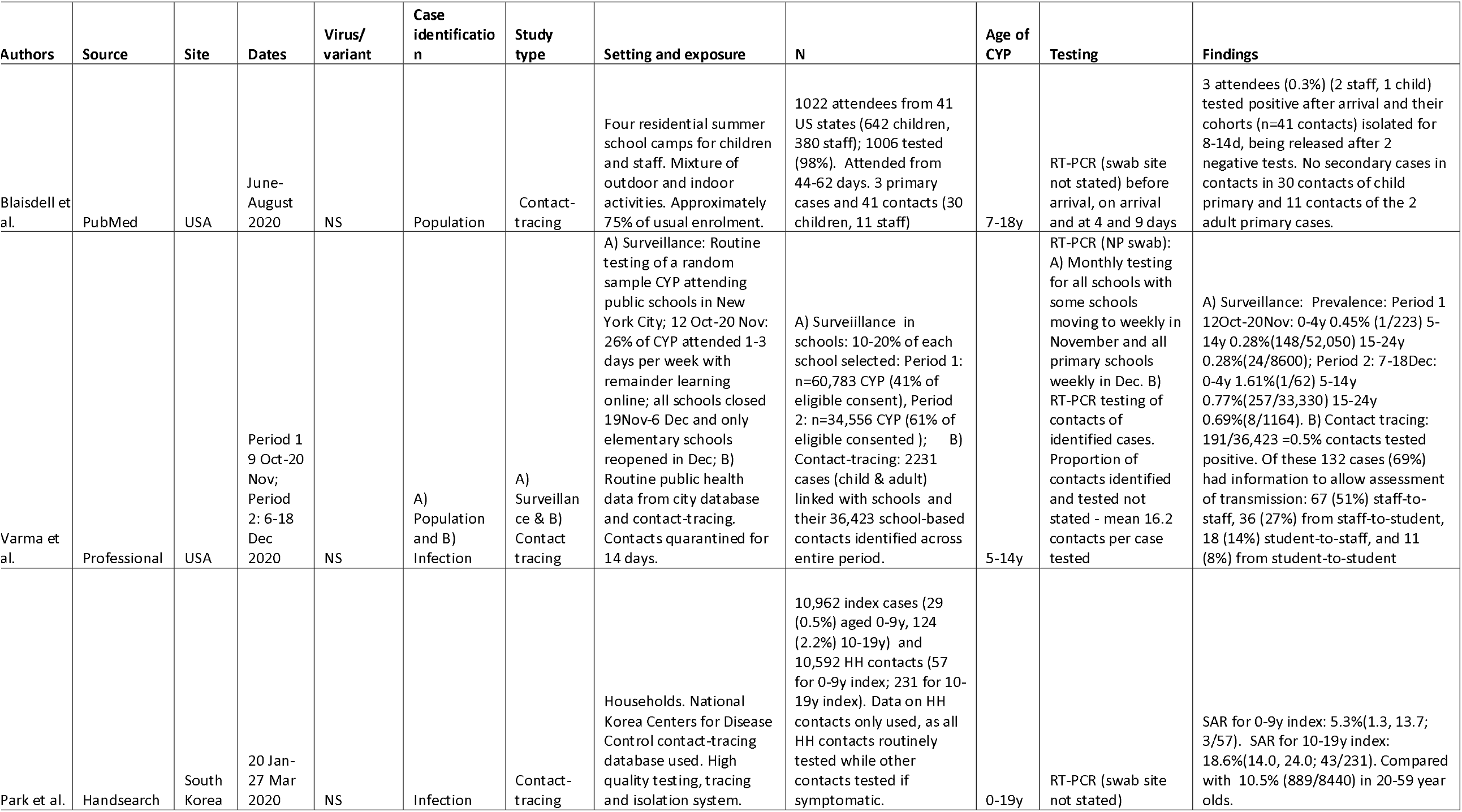
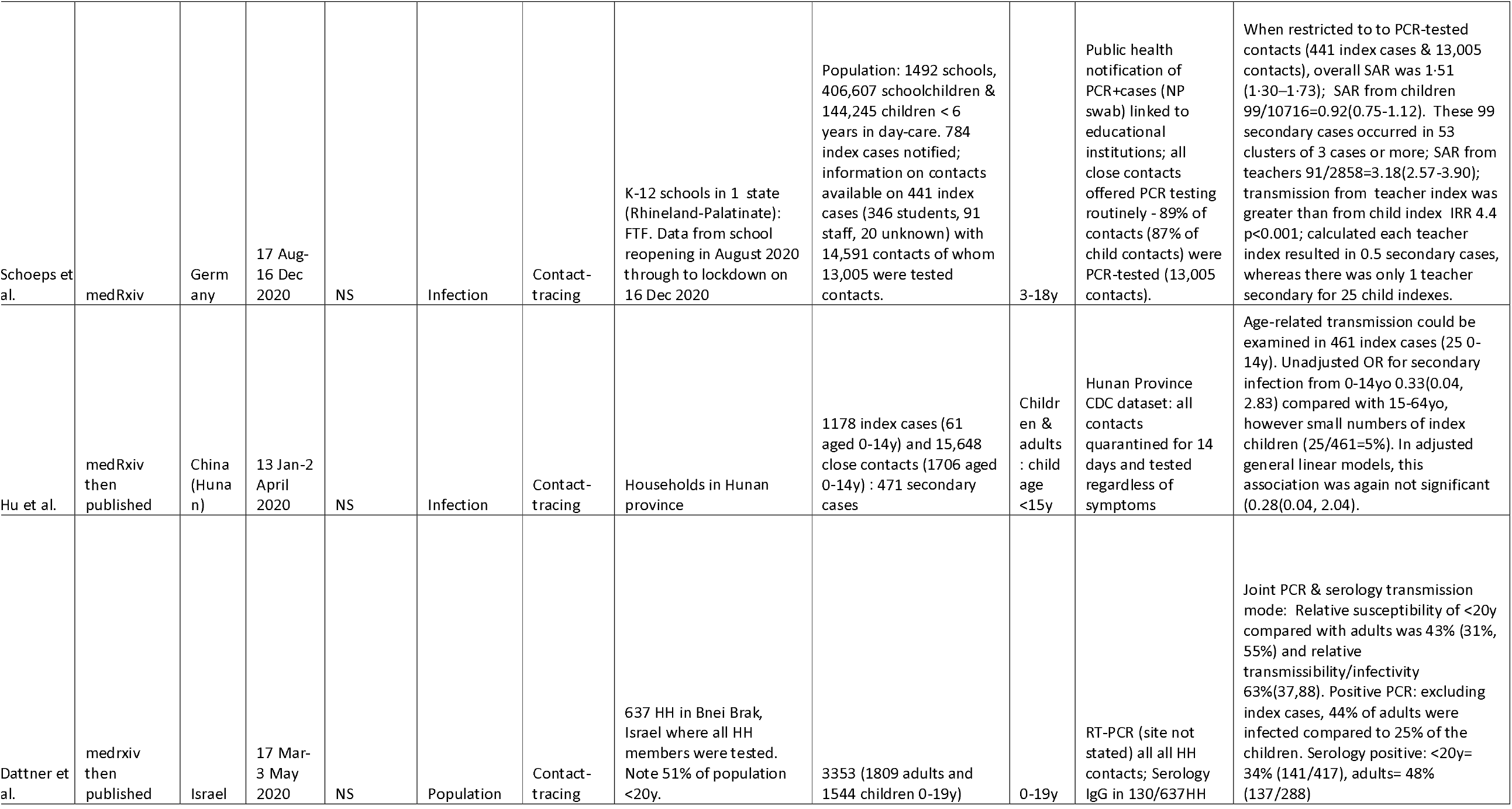
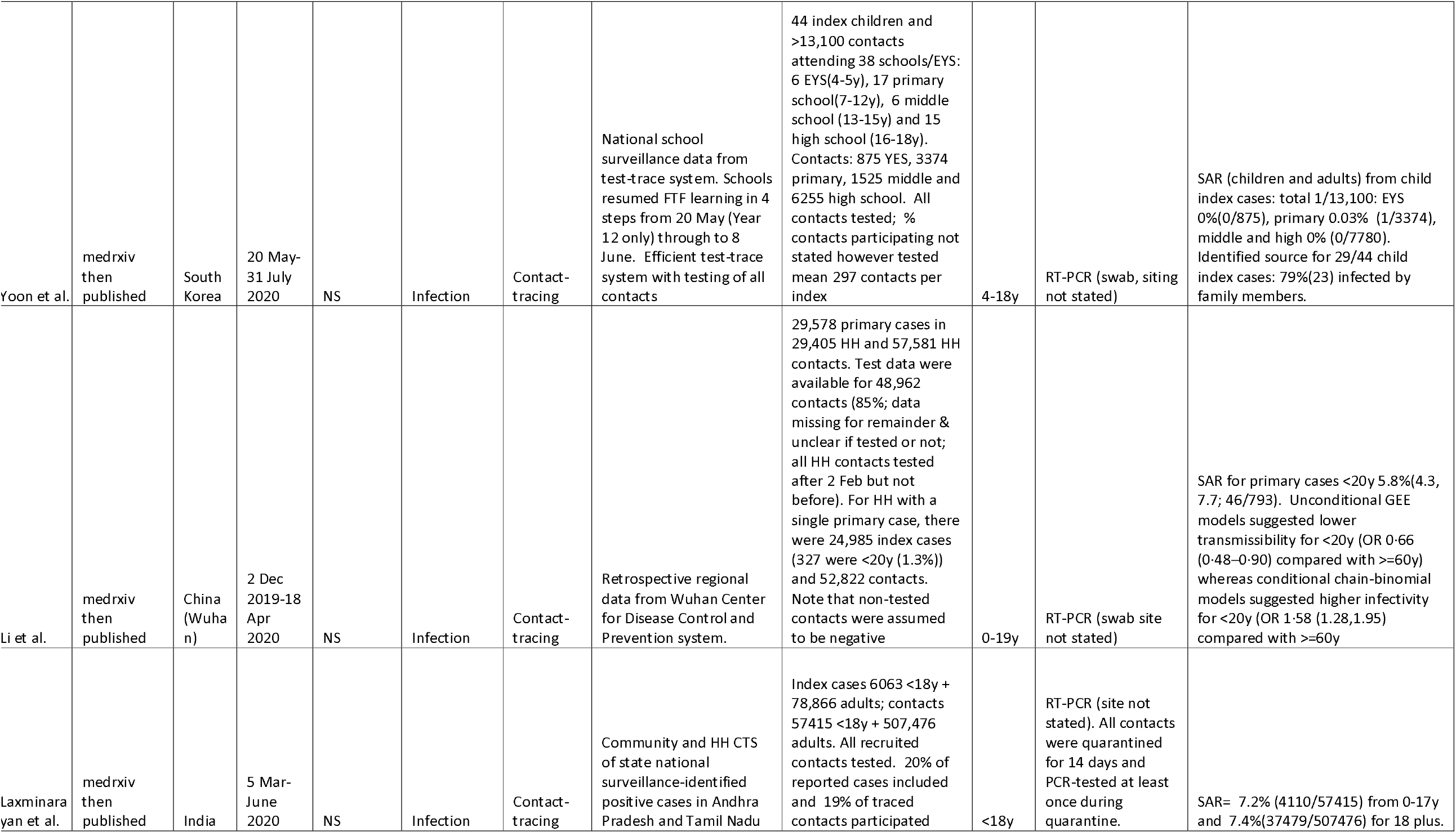
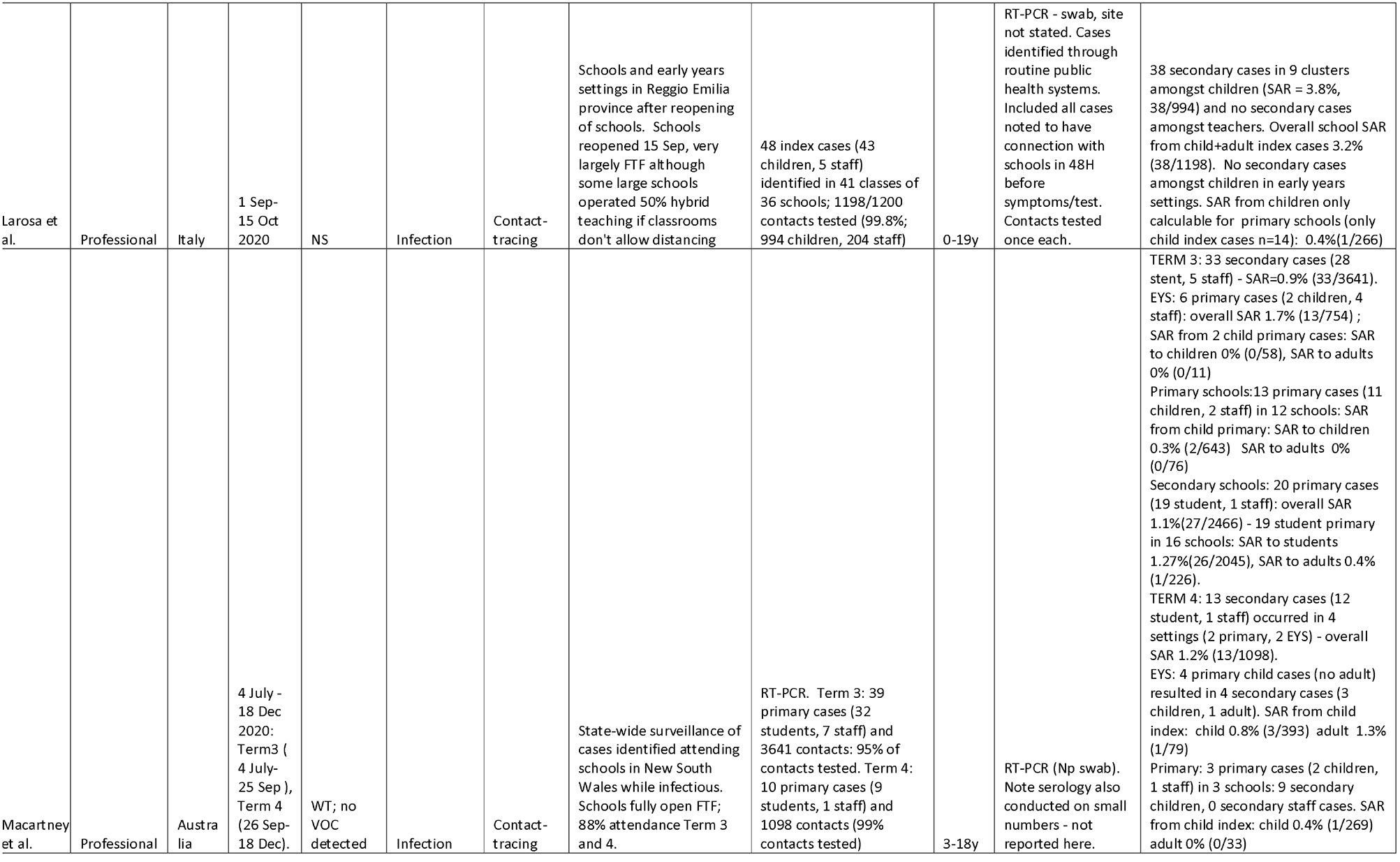
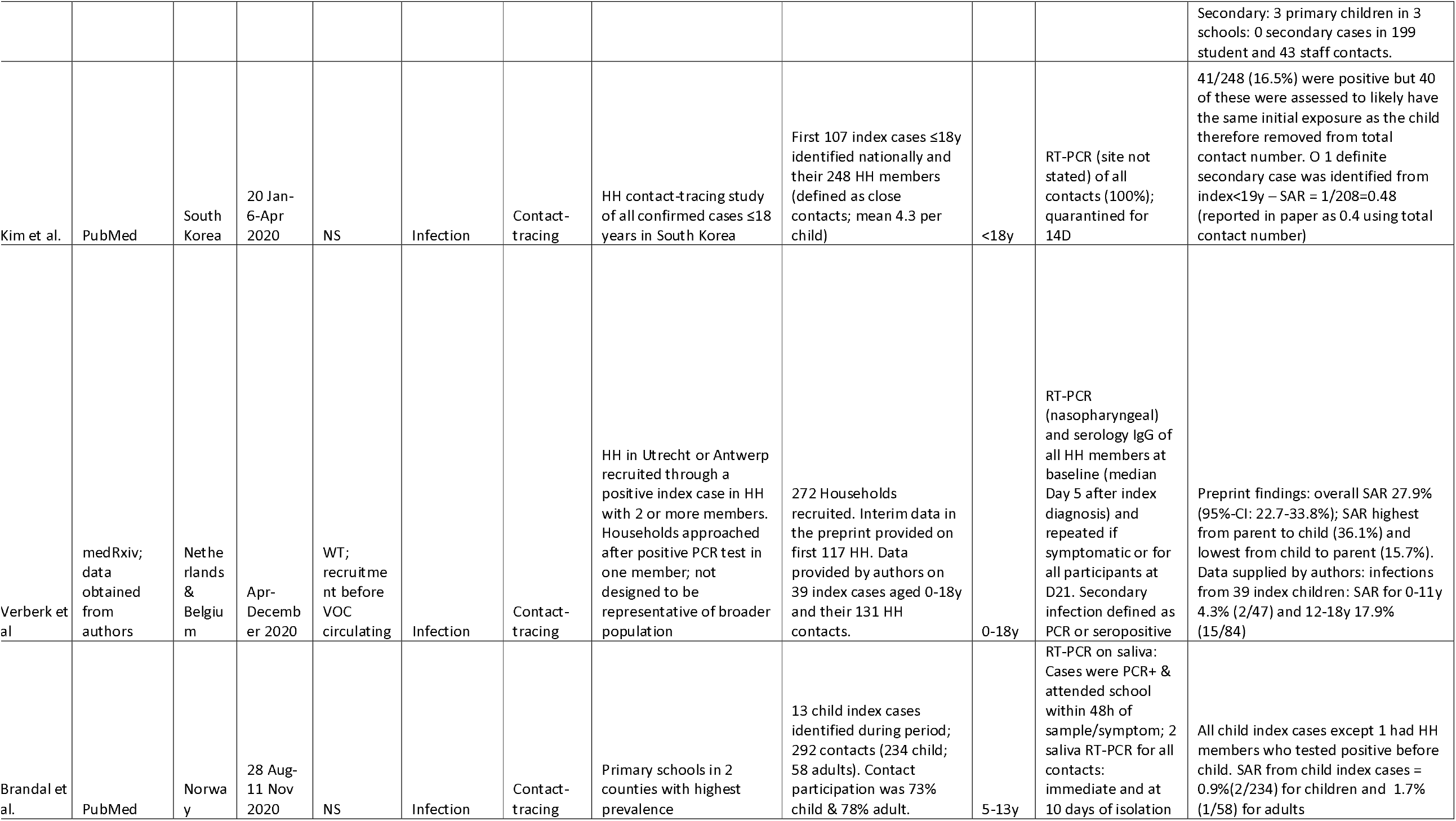
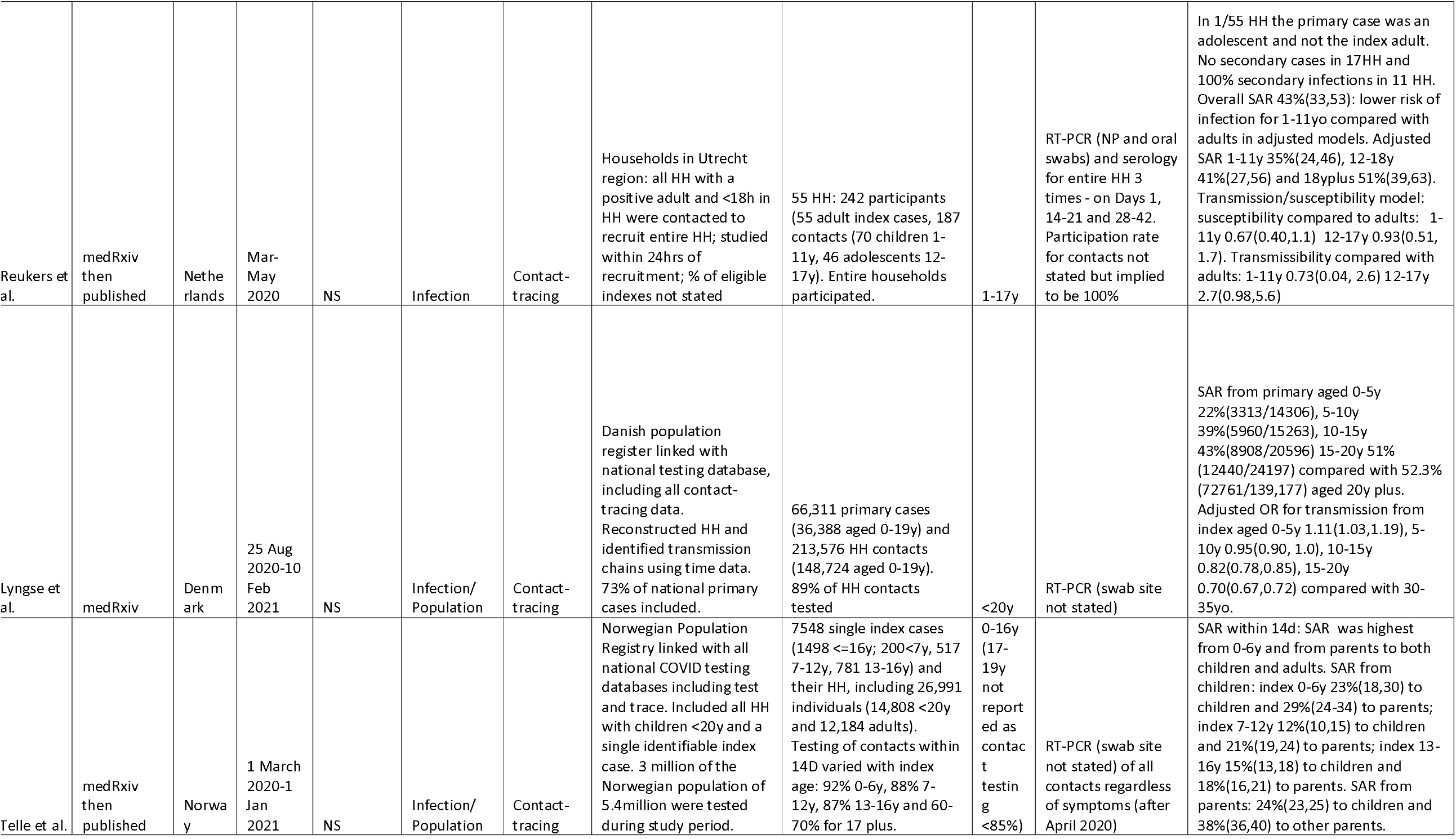
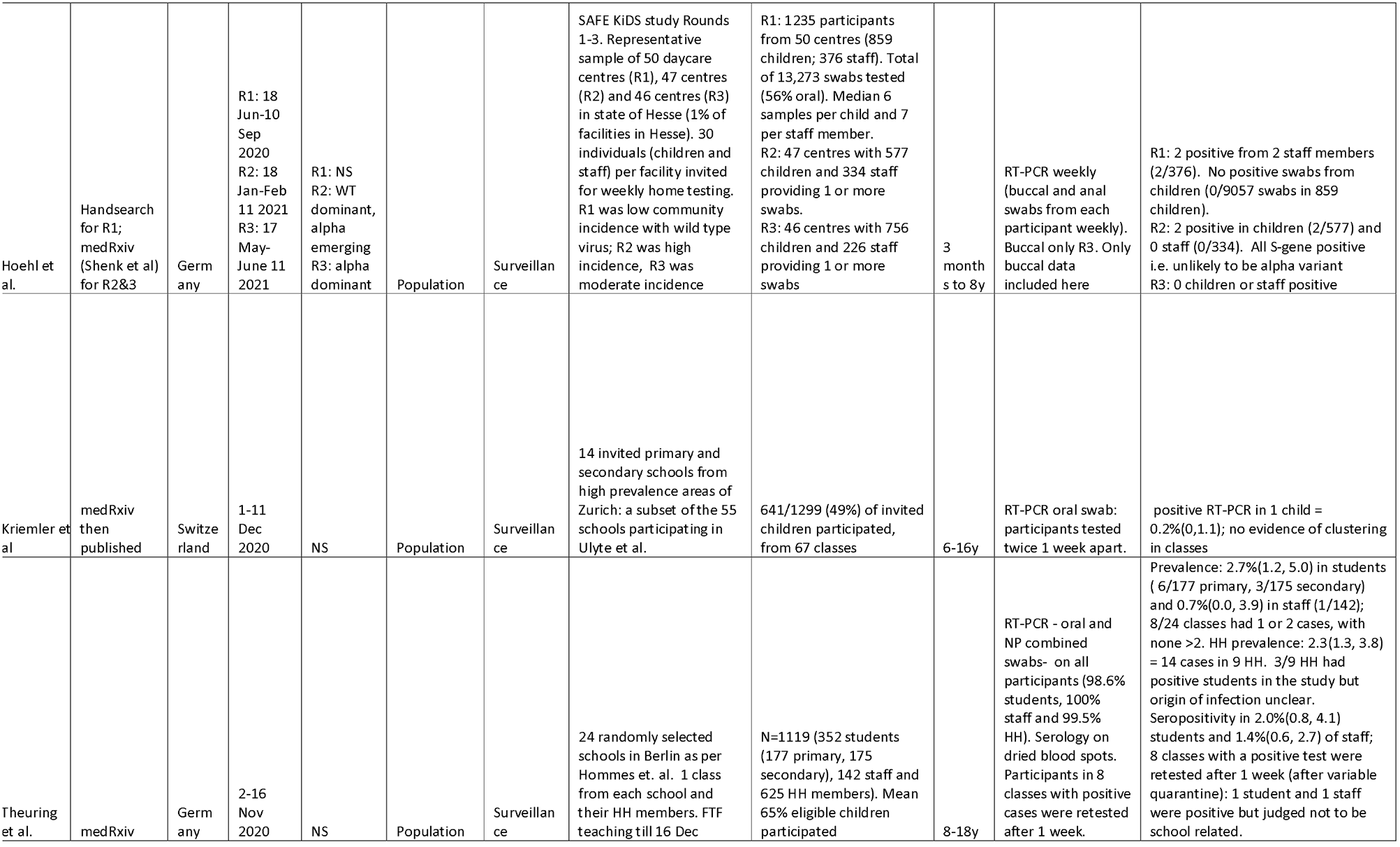
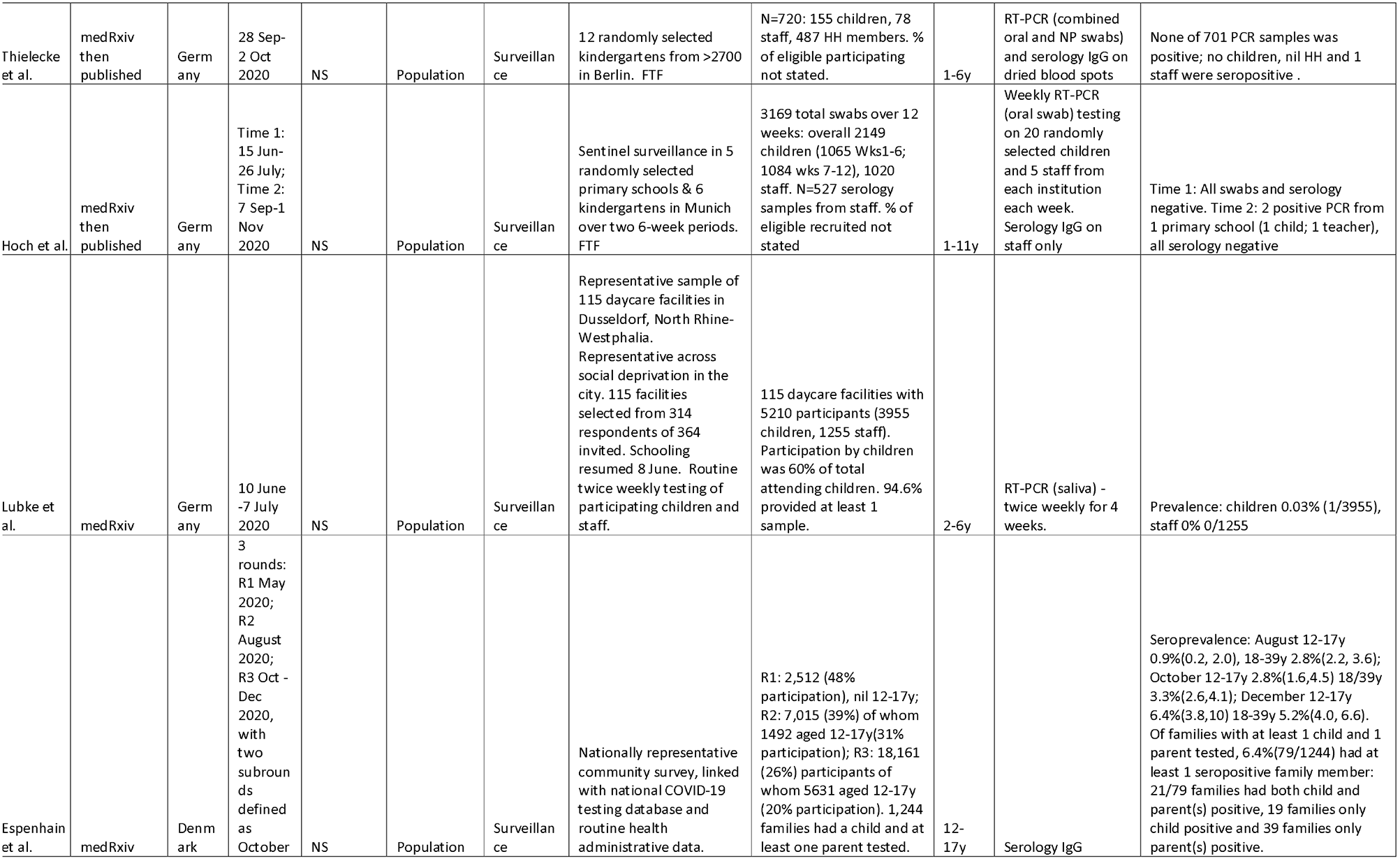
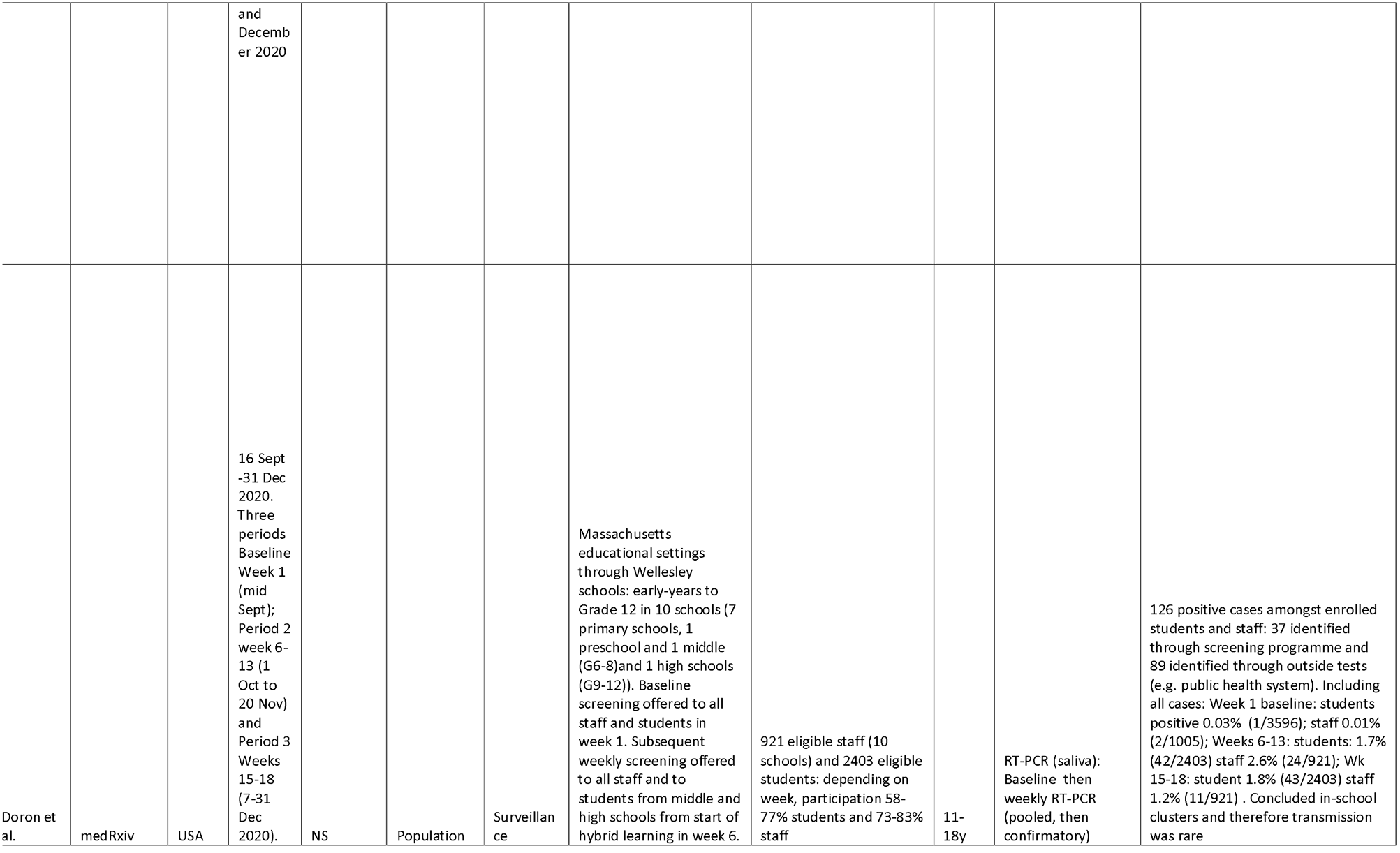
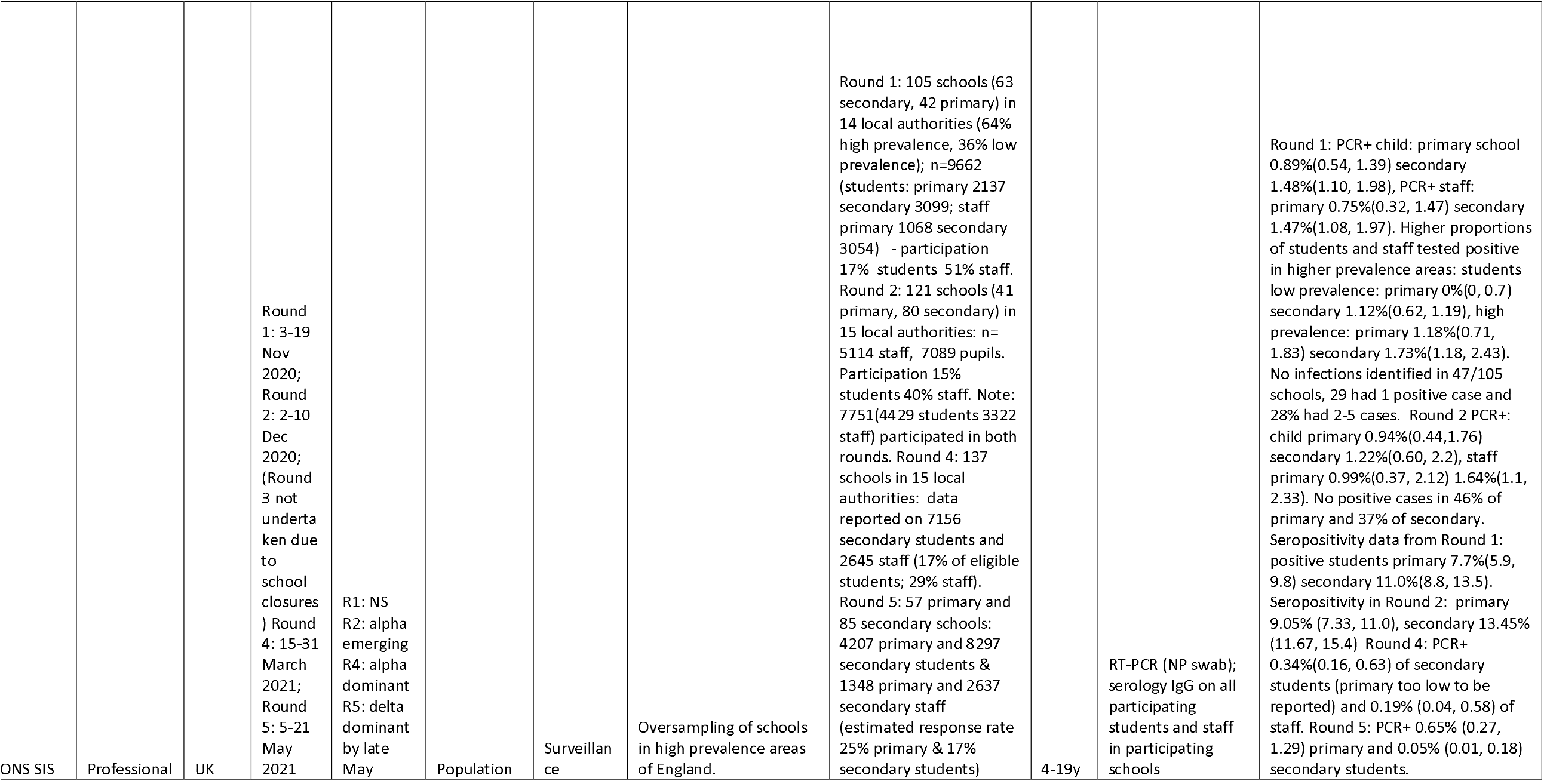
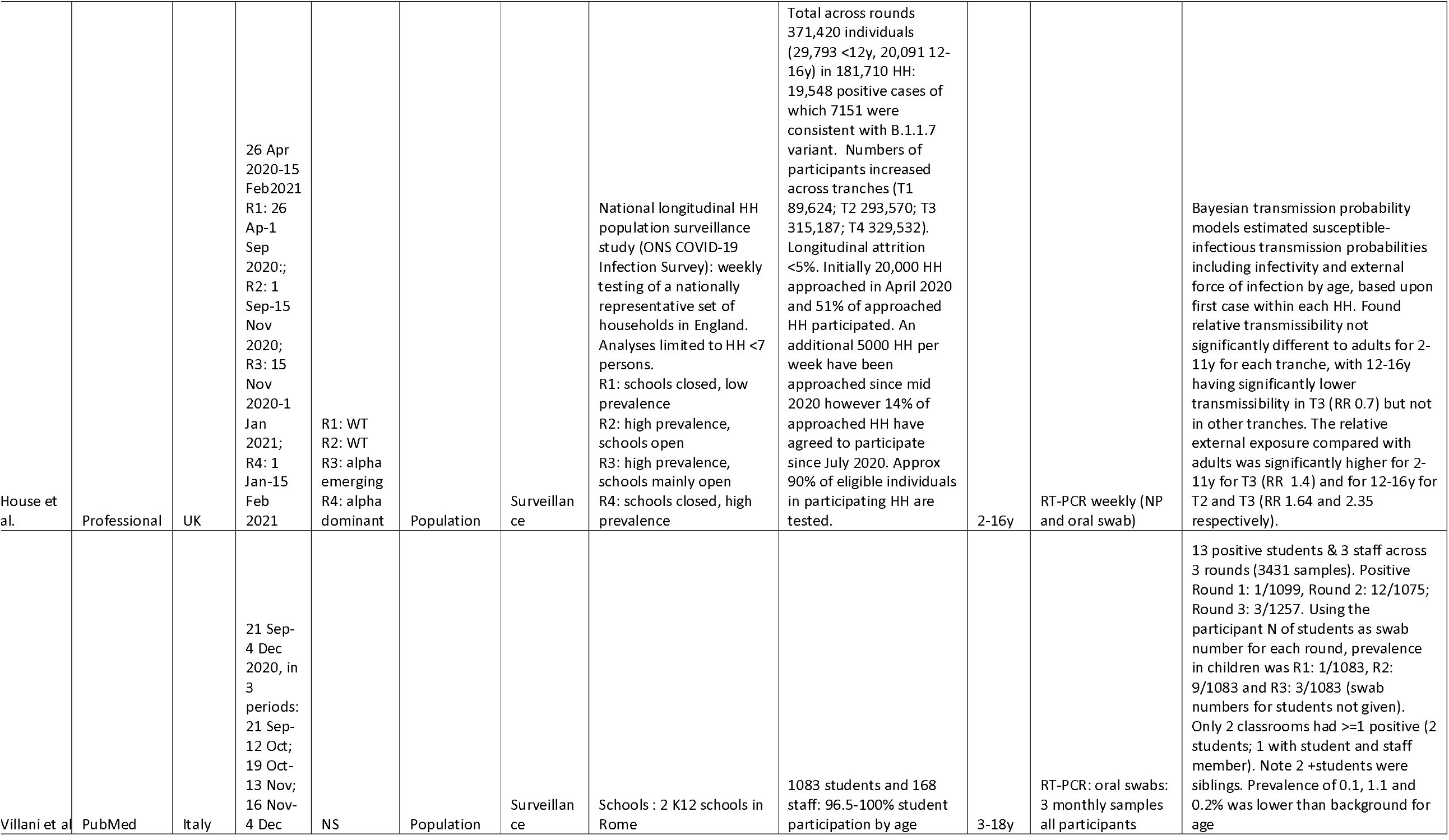
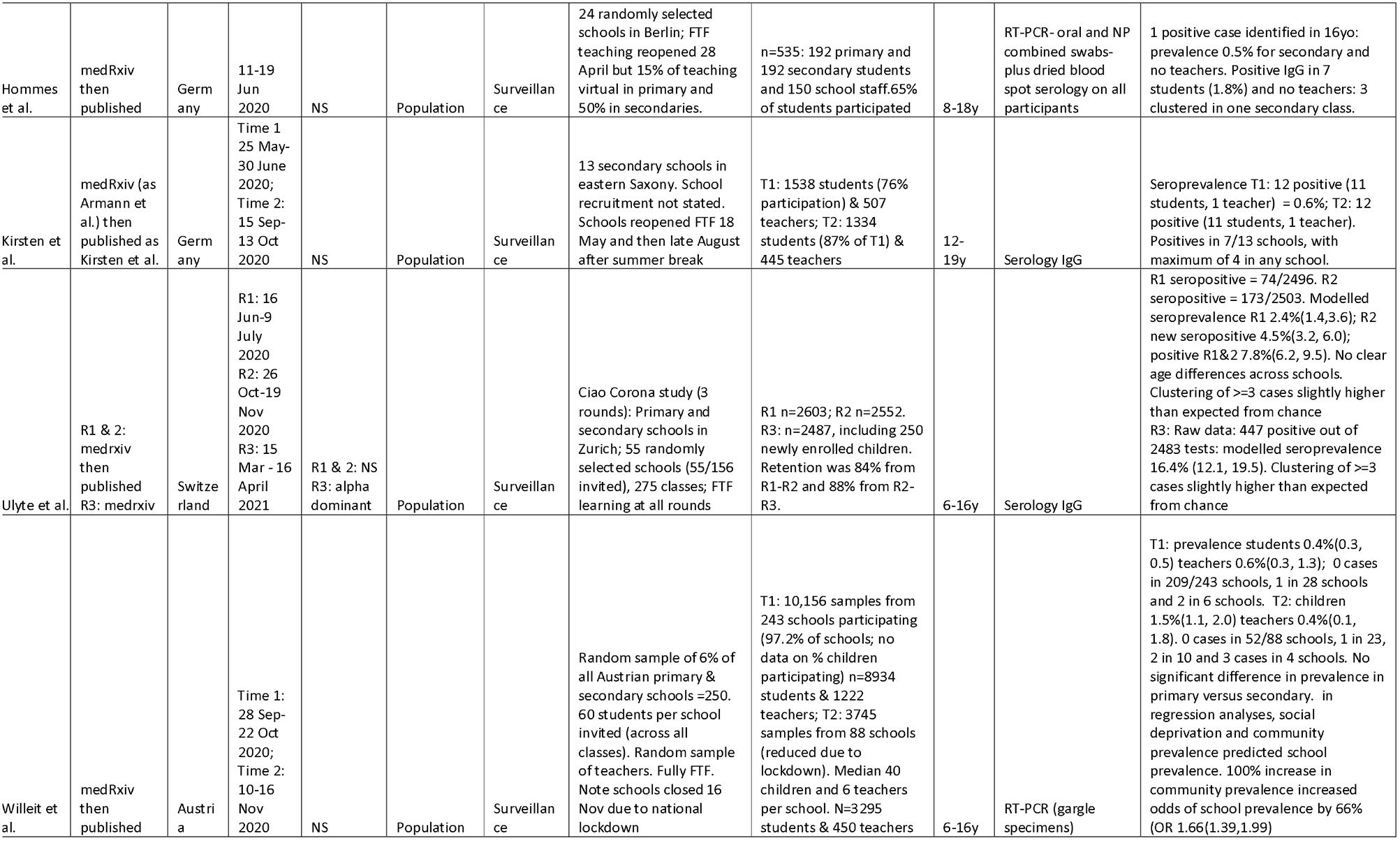
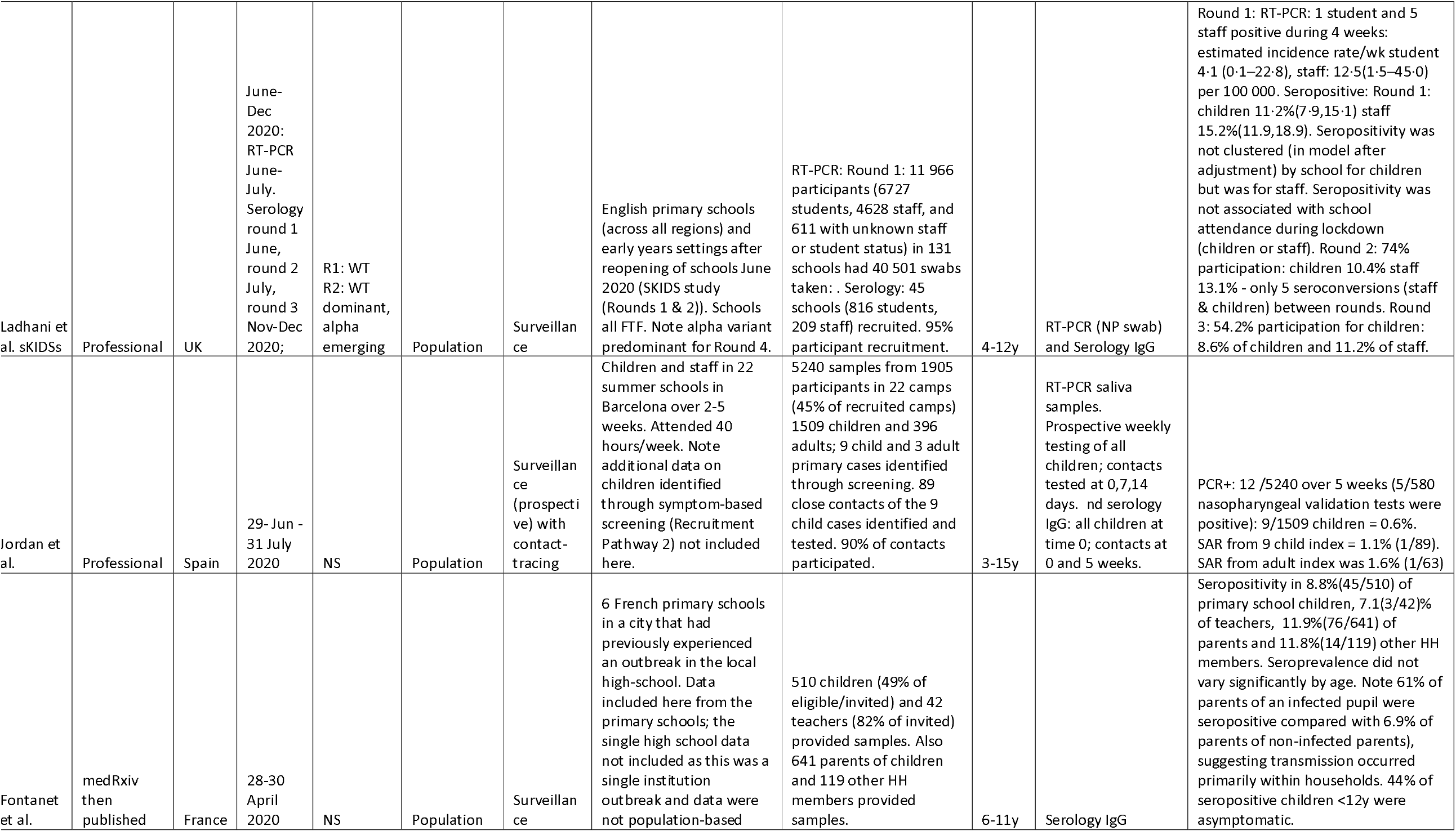
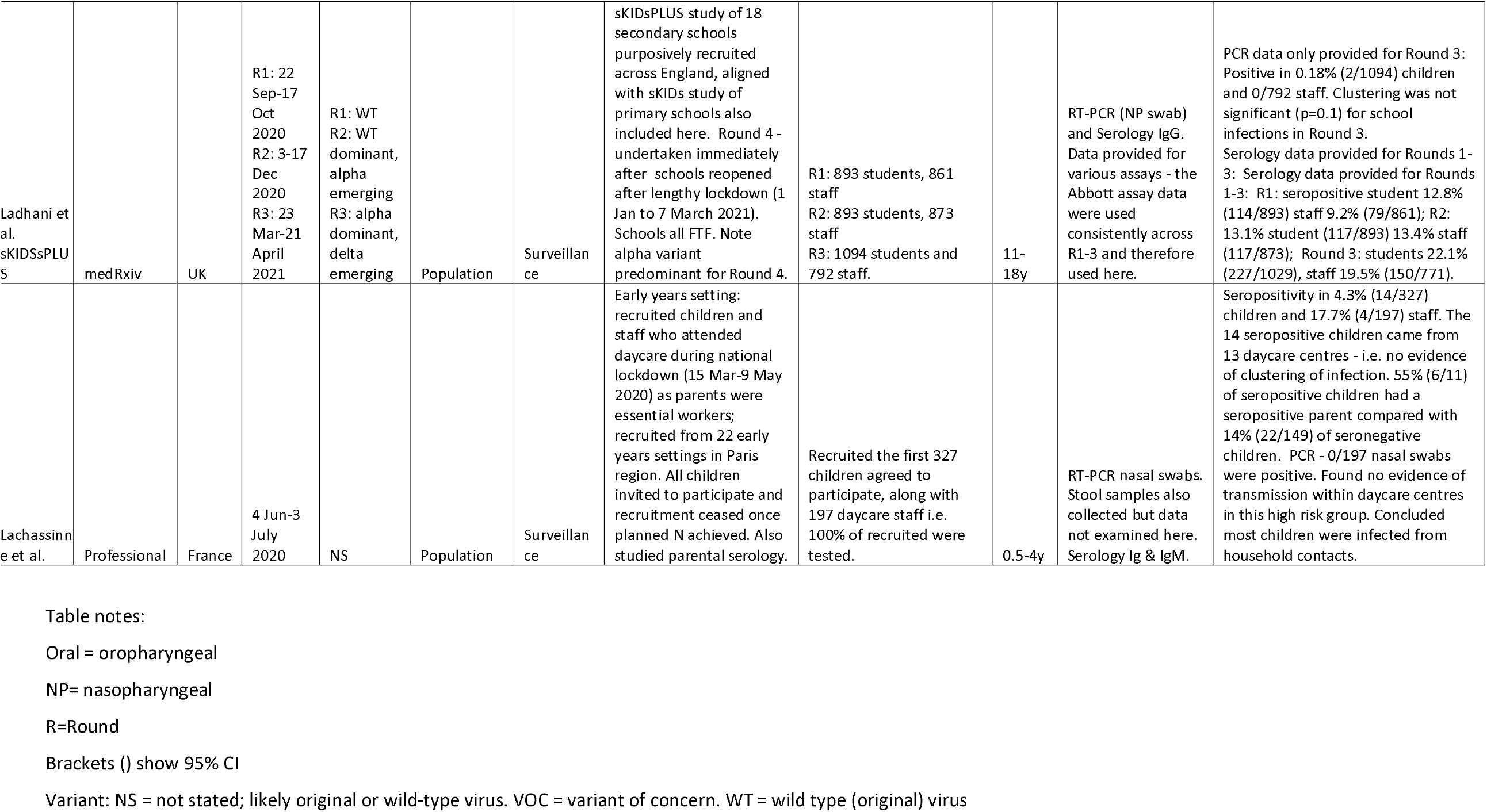
Study characteristics

Sixteen studies were contact-tracing studies (6 school;[27-33] 10 household[2, 34-42]), 2 provided both contact-tracing and population data (both school studies[43, 44]) and 19 were population studies (17 in educational settings;[9, 10, 45-59] 2 were national community surveillance surveys[60, 61]).

Twenty-four studies were high quality (13 population; 10 contact-tracing and 1 study providing both data) and 13 studies were medium quality (6 population, 6 contact-tracing and 1 study providing both data). Of the 43 articles reporting the 37 studies, 26 (60%) were published, 11 (26%) were preprints and 6 (14%) were government or university reports.

Eight studies were from Germany, 4 from the UK, 3 from South Korea and the USA, 2 each from China, France, Switzerland, Denmark, Italy and Norway, one included data from both the Netherlands and Belgium, and 1 study each from Netherlands, Austria, Israel, India, Spain, and Australia.

Thirty-one studies (84%) were undertaken before November 2020 and involved the wild-type virus, although only 2 explicitly reported this; 6 (16%) studies included rounds with the alpha variant emerging (1) or dominant (5), with 2 (5%) also including rounds in which the delta variant was emerging.

### Contact-tracing studies (household and school)

Eighteen studies provided data on secondary infection or attack rates (SAR) from child index cases, including five large regional[2, 31, 32, 35, 37] and five national[34, 38, 41, 62, 63] studies. Fifteen (8 household;[2, 34, 35, 37-39, 41, 63] 7 school[27-33, 44]) provided sufficient data to include in meta-analyses of secondary attack rates.

Forest plots of SAR from child index cases to all-age contacts are shown in Figure 2 separately by setting. The pooled estimates of SAR were 7.6% (3.6, 15.9) for household studies (panel A), significantly higher than the pooled estimate for school studies of 0.7% (0.2, 2.7) (panel B) (difference QM (df=1) = 9.325, p=0.0023).

**Figure 2.**
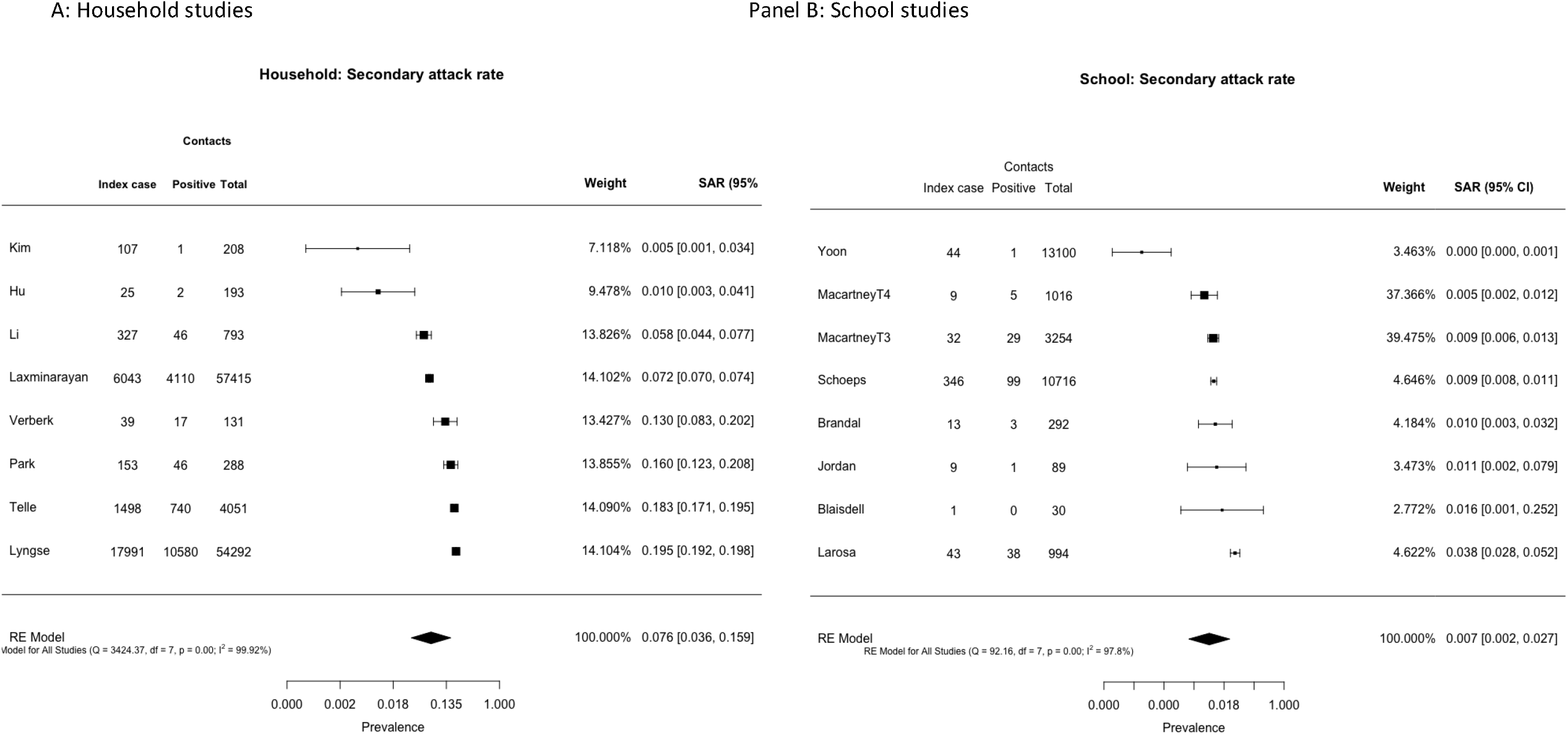
Secondary attack rates from child index cases to all contacts for (A) household studies and (B) school contact-tracing studies

Transmission from child index cases by age of contacts could be assessed in 4 school studies and 1 household study (Appendix Figure 1). Pooled SAR to child contacts was not different to that to adult contacts (p=0.45).

Odds of being a secondary case (of any age) from a child index compared with an adult index case were calculated from 11 rounds of data (6 household, 5 school; see Figure 3). Across all studies, pooled risk of transmission was lower from child index cases than adults (OR 0.49 (0.25, 0.98); in sub-group analyses the OR was 0.27 (0.06,1.28) for school studies and 0.72 (0.45, 1.16) for household studies, all with high heterogeneity.

**Figure 3.**
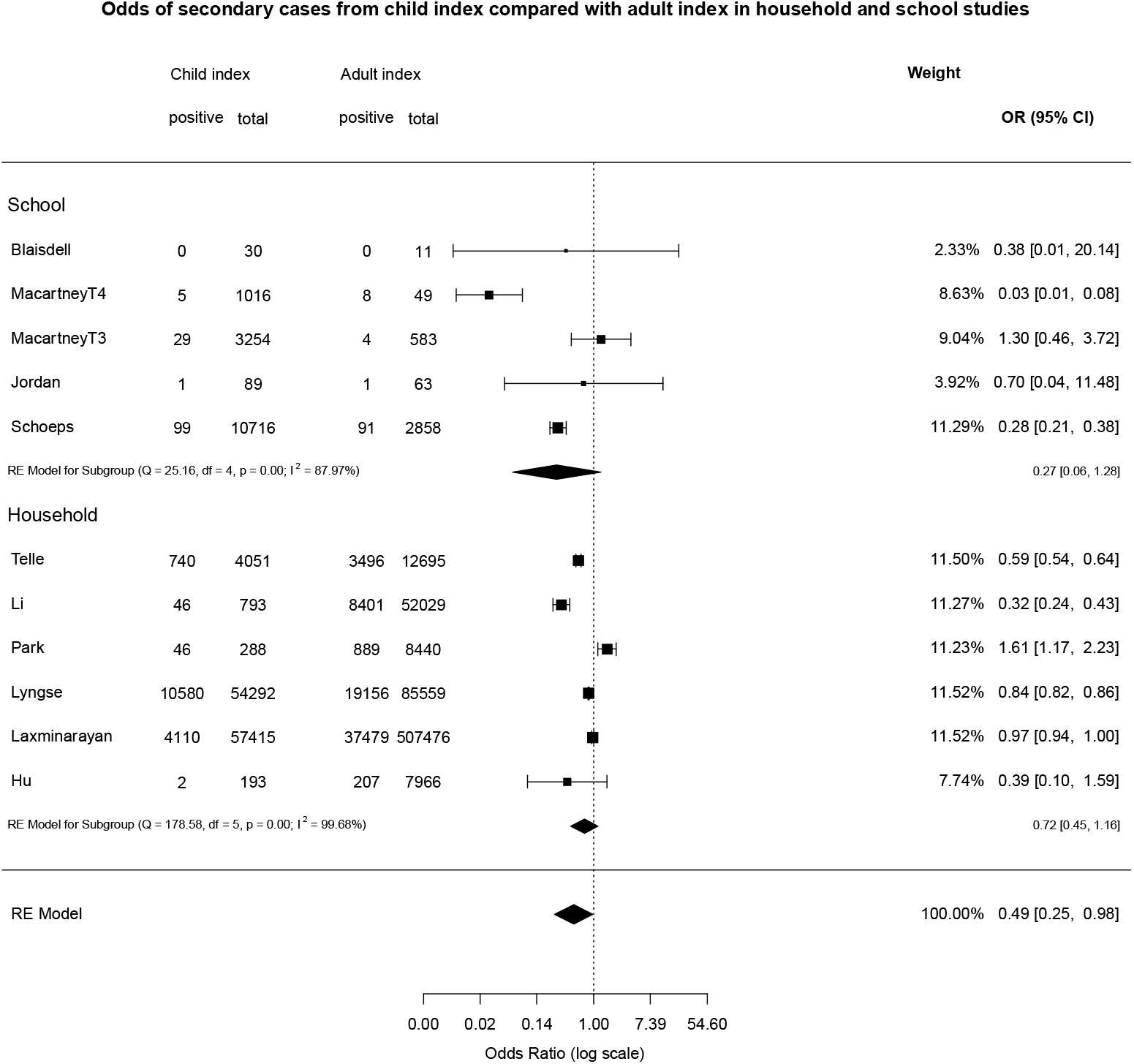
Odds of being a secondary case from child compared with adult index cases

Two studies could not be included in the meta-analyses. Varma et al. undertook a large school contact-tracing study from New York City[43] and reported that the overall school SAR from CYP and adults was 0.5%; of the 69% of secondary cases for which a source of infection could be identified, 51% were staff-to-staff, 27% staff-to-student, 14% student-to-staff, and 8% from student-to-student. Espenhain et al.[61] used data from 4 rounds of a Danish nationally representative community survey to examine transmission in 1244 households with resident adolescents. They reported that, in 73% of families with at least one seropositive family member, only the parent(s) or the child were seropositive, concluding that transmission between generations was uncommon.

### Adjusted household transmission models

Six studies examined transmission from CYP to household members using adjusted transmission models accounting for a range of factors including individual exposure histories, potential tertiary transmission, poverty and the age-structure of populations. Two studies used nationally representative data from England[60] and Denmark,[41] and four were contact-tracing studies (from China,[35, 37] Israel[36] and the Netherlands[40]).

House et al.[60] used longitudinal weekly PCR testing from a very large representative national sample of English household[64] to estimate susceptible-infectious transmission probabilities from models in four periods from April 2020 to February 2021 across low and high prevalence, schools being reopened and the emergence of the alpha (B.1.1.7) variant in late 2020. They found transmissibility did not differ by age. However they did observe that the risk of bringing infection into household (relative external exposure) was higher amongst 12-16y than for adults although these included periods of national lockdown for adults whilst all children continued to attend full-time schooling. A Dutch contact-tracing study similarly concluded there were no differences in transmissibility between children and adults,[40] whilst a large national Danish study[41] and an Israeli contact-tracing study[36] found lower relative transmissibility in children and young people compared to adults. Two contact-tracing studies from China found that, whilst in unadjusted analyses infected children generated fewer secondary cases than adults, adjusted models showed no difference,[35] or higher infectivity.[37]

Multilevel random-effects meta-analysis of relative transmissibility from CYP compared with adults included 13 estimates from 6 studies with total person-observations from 127,822 CYP and 1,526,117 adults (Figure 4). The pooled relative transmissibility from CYP was 0.92 (0.68, 1.26) compared with adults, with high heterogeneity (99.43%). Data did not allow sub-group analyses by age of child.

**Figure 4.**
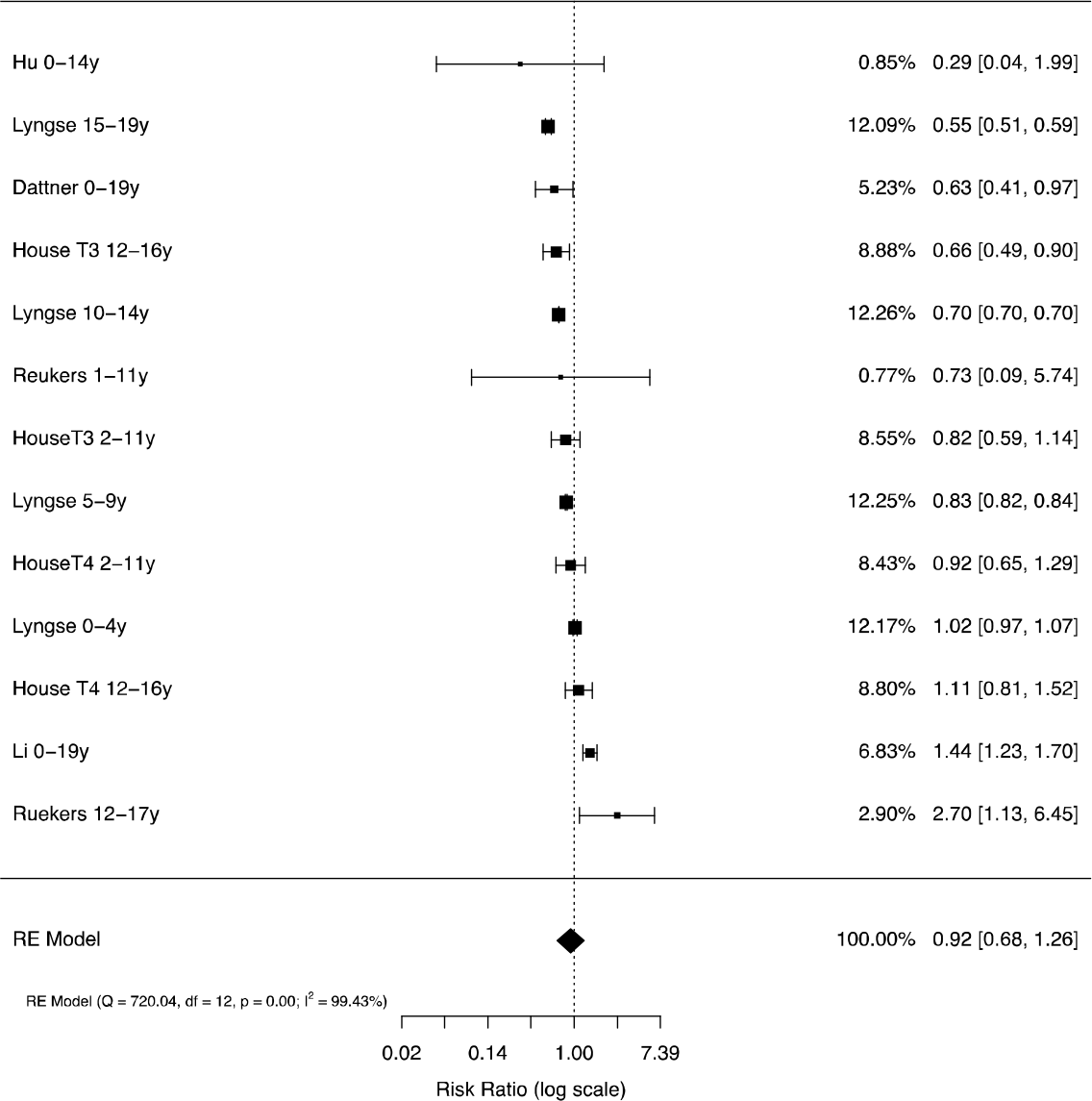
Relative transmissibility of children and adolescents compared with adults in adjusted household models Note: Analysis includes the last two periods from House et al. and estimates by age from other studies.

### School prevalence studies

Infection prevalence in schools or nurseries was measured in 16 studies (31 rounds of observations; total 161,280 child-observations) and antibody prevalence was measured in 9 studies (20 rounds; 26,509 child-observations). Some provided data for single age-groups (e.g. early-years, primary or secondary students) while others provided cross age-group data. In the main analyses, we used overall estimates where they exist and estimates by age-group where the former were not provided.

Forest plots of PCR prevalence and seroprevalence by age are shown in Figure 5. Meta-regression models are shown in Table 2. Pooled infection (PCR) prevalence across all studies was 0.4% (0.2, 0.6), not significantly different by age-group (p=0.32). Prevalence was also associated with contemporary community 14-day incidence (OR 1.003 (1.001, 1.004), p<0.001) and prevalence in the month prior to the study (OR 1.003 (1.001, 1.006), p=0.008) but not with 2 months prior. PCR prevalence was not associated with school attendance rate or PCR source. Plot of predicted school prevalence by 14-day incidence is shown across age-groups in Figure 6.

**Figure 5.**
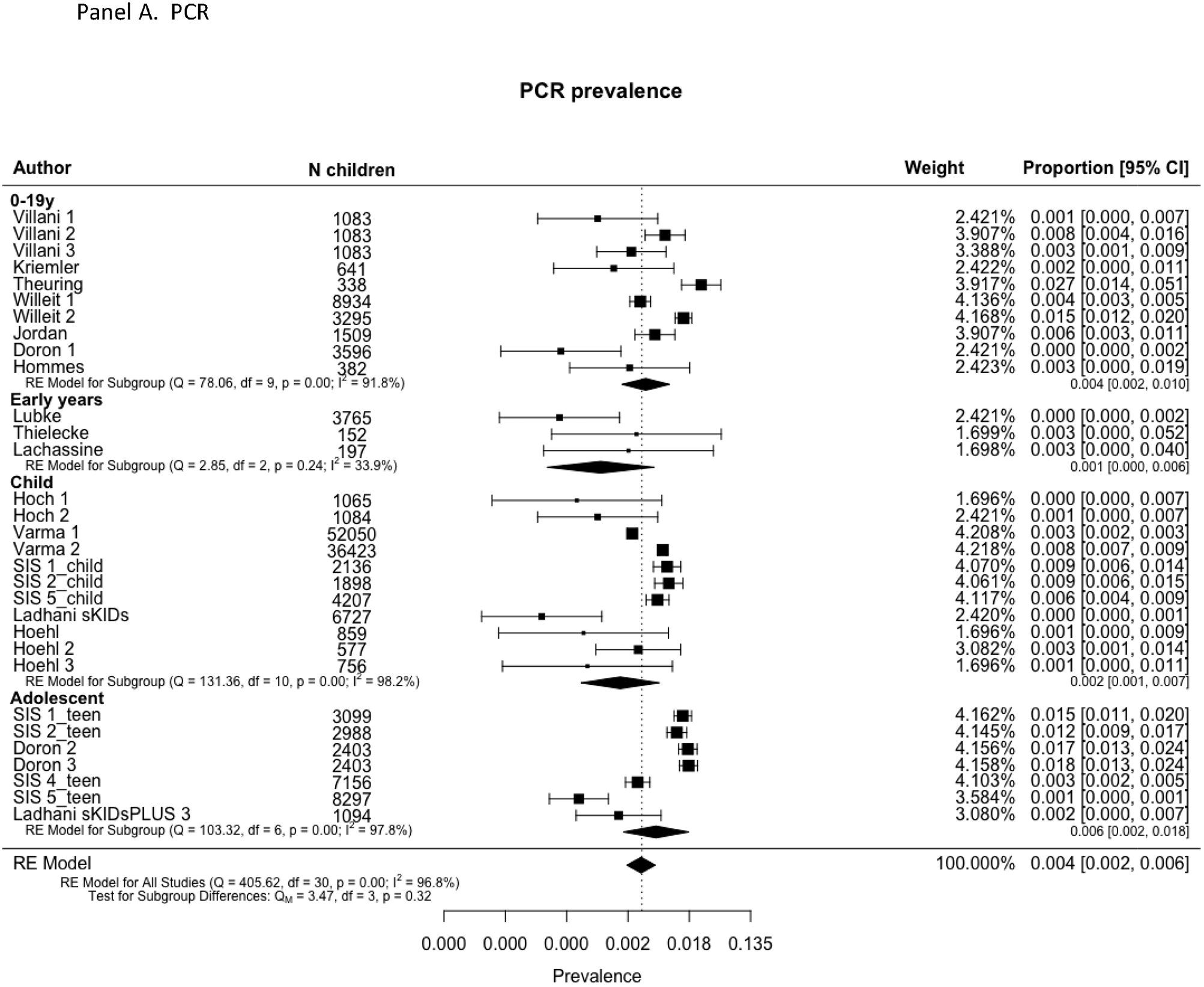
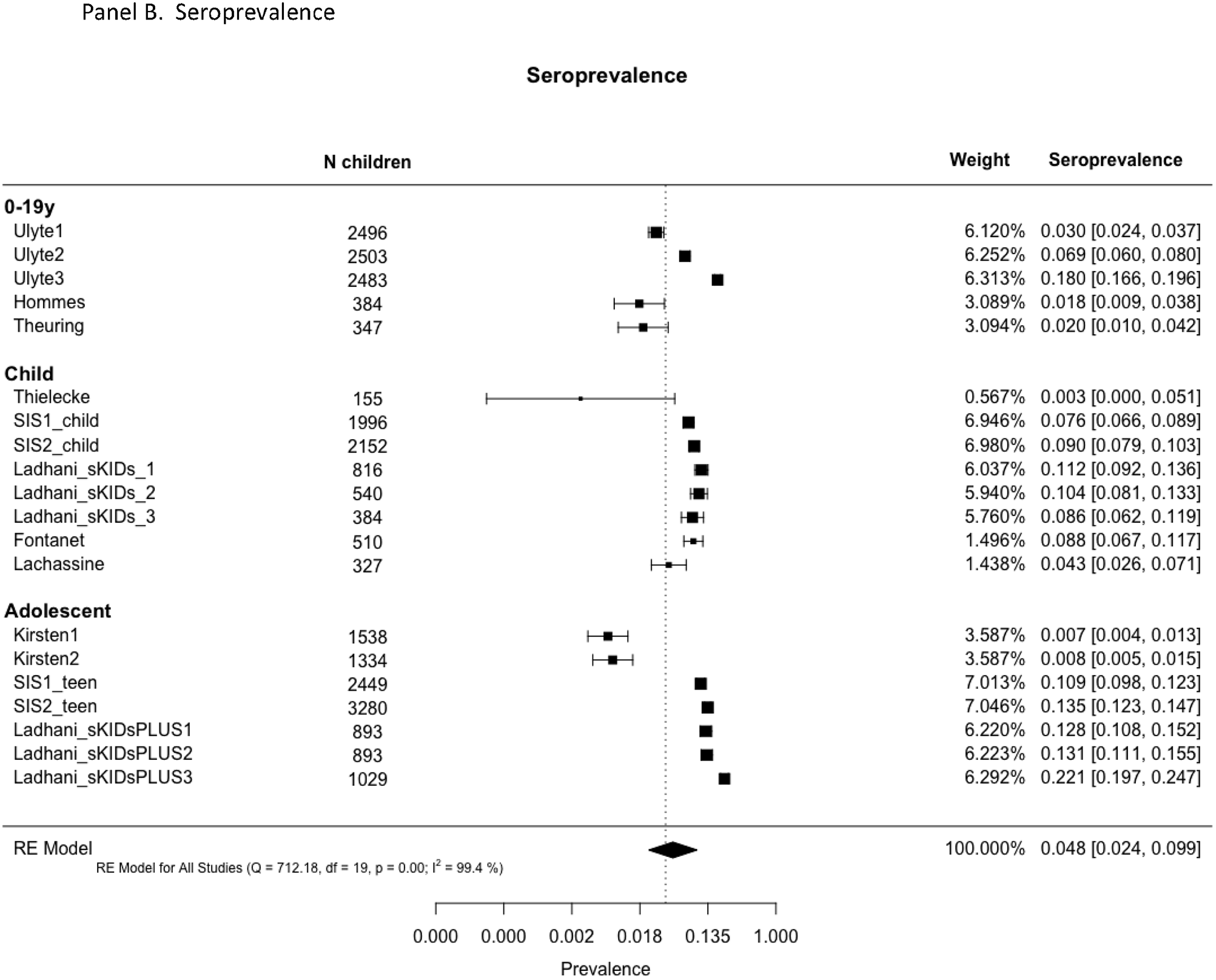
Prevalence and seroprevalence of SARS-CoV-2 infection in schools by age-group: (A) PCR prevalence and (B) Seroprevalence.

**Figure 6.**
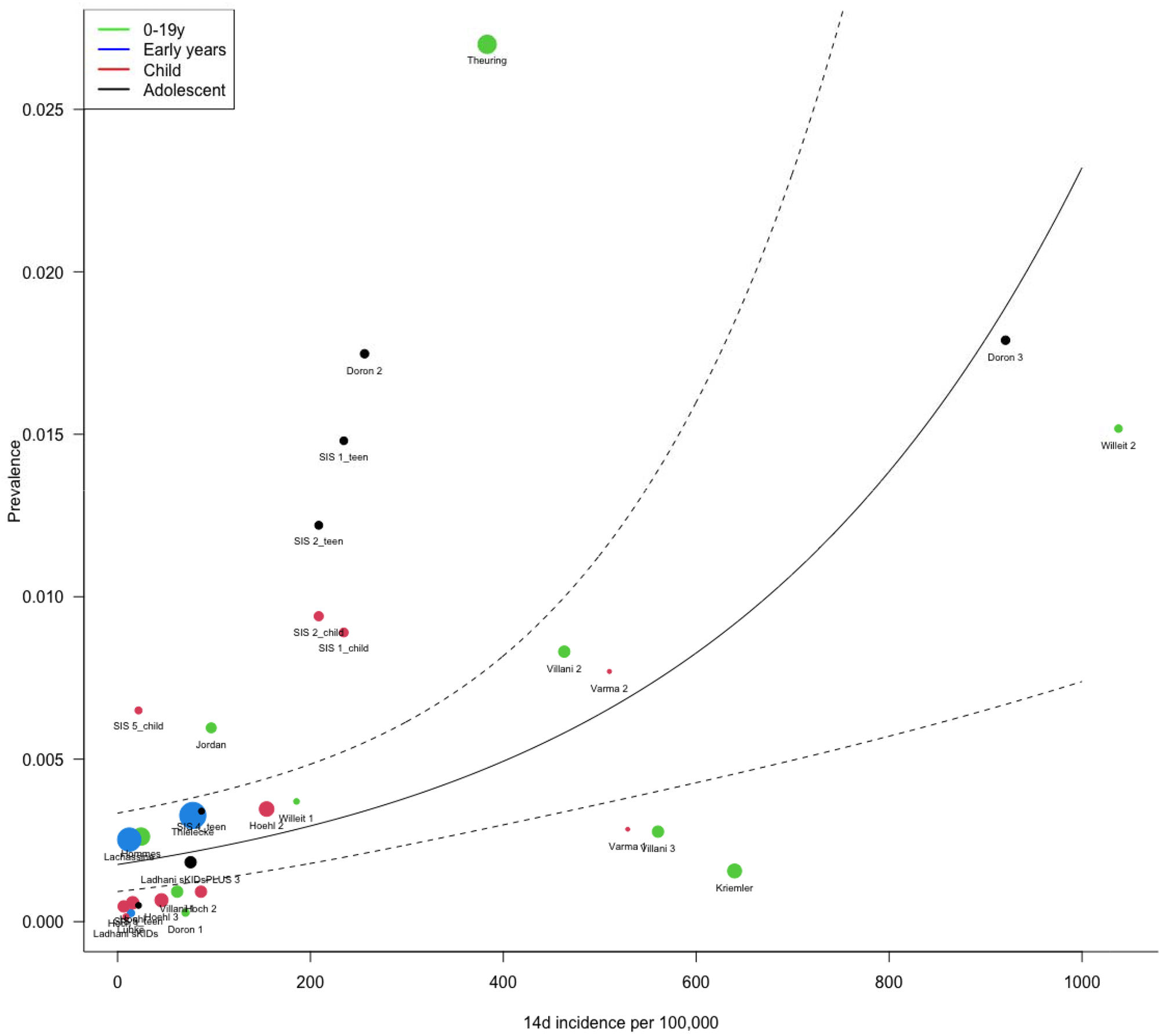
Plot of predicted prevalence and 95% CI in school studies by community 14-day incidence of SARS-CoV-2 infections per 100,000

**Table 2.**
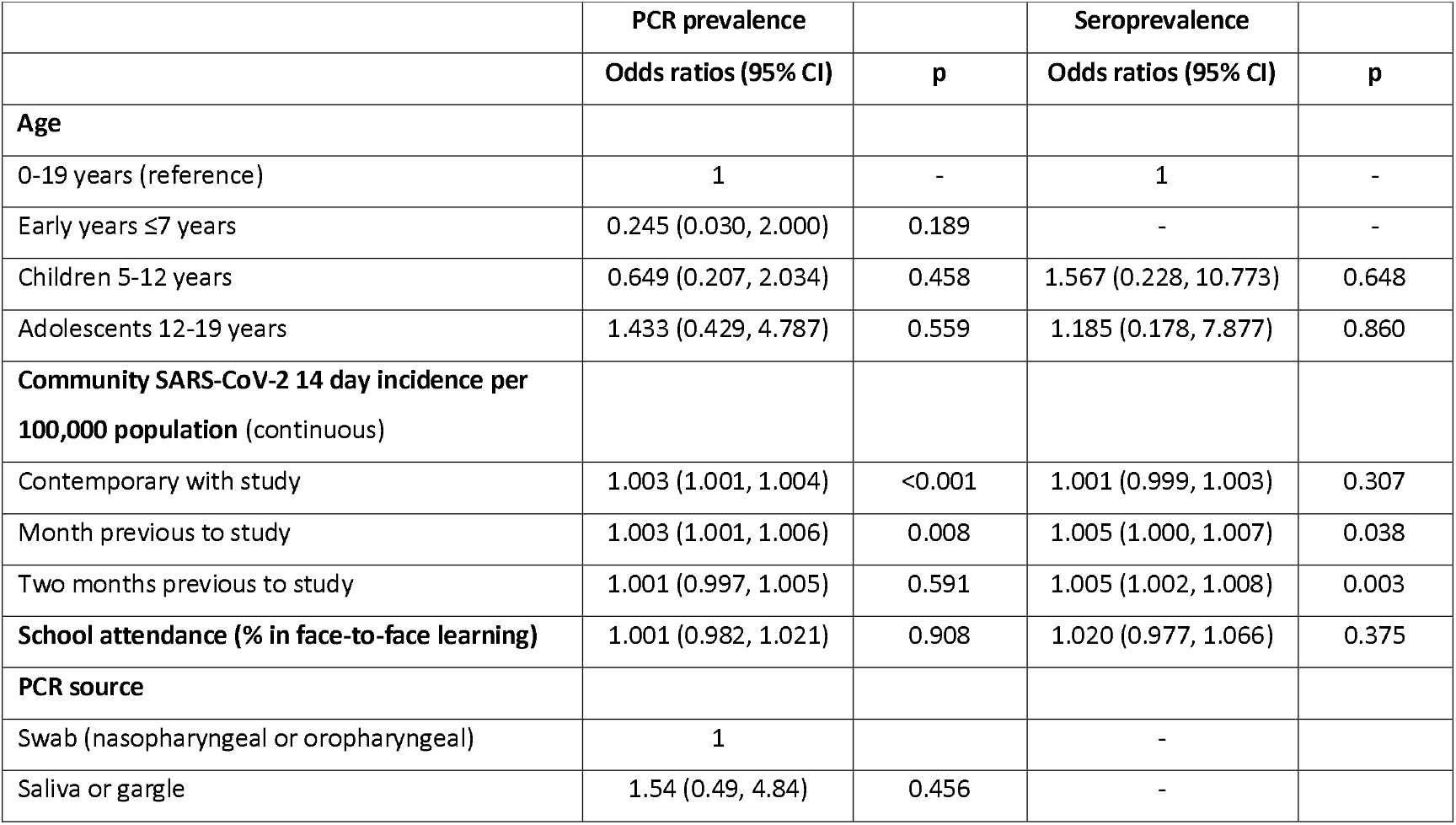
Moderators of prevalence and seroprevalence in school studies

Pooled seroprevalence across all studies was 4.8% (2.4, 9.9), with no significant difference by age-group. Seroprevalence was associated with community incidence in the month and two months prior to the study, but not with contemporary incidence. Seroprevalence was not associated with school attendance.

No school studies fitted adjusted transmission models. Only two studies undertook a detailed analysis of clustering; Ulyte et al.[9, 65] reported that clusters of ≥3 cases occurred in 7 of 129 classes in Round 2 and 24 of 119 in Round, more than the 4 and 17 classes expected by chance respectively. A very large school contact-tracing study by Schoeps et al.[28] reported that 83% of 784 school index cases led to no secondary cases. All other studies reported no evidence of clustering of infections (i.e. > 3-5 infections per class) within schools.[10, 46, 47, 51-56, 59, 66, 67] Other observations supporting limited transmission in schools were calculations showing that where direction of transmission was available, the majority appeared to be from adults to children[28, 43, 49, 51, 68] or that origins of transmission chains were outside schools;[47] and observations that virus prevalence in school children and teachers was lower than in the local community at the time despite higher levels of testing within schools.[43, 52, 53, 67] Seroprevalence studies, however, reported similar antibody prevalence amongst students and teachers[54, 67, 68] or adults in the local community.[9, 67, 68]

The association of school prevalence with community infection rates was examined in two school studies, both of which reported positive associations.[43, 56] Only one study examined associations of prevalence with social deprivation, reporting a positive association.[56]

## Discussion

We report the first findings relating to SARS-CoV-2 transmission from CYP through meta-analysis of studies with low risk of bias. Meta-analysis of household studies which undertook adjusted transmission analyses showed no difference in relative transmissibility between CYP and adults (OR 0.92 (0.68, 1.26)), although meta-analysis of unadjusted secondary attack rates suggested that transmission from CYP was lower than from adults, although with wide confidence intervals. There are a number of sources of potential bias in the unadjusted analyses, including low numbers of child index cases as well as differential transmission from children across generations of spread within households, and it is likely that these analyses under-estimate relative transmissibility. These findings suggest that, within households, CYP play a role in transmission that is to similar but not higher than adults. The only study to examine external force of infection suggests CYP play a role in bringing infection into the house when schools are open, but this included periods when the country was in lockdown whilst schools remained fully open.[60]

We found a striking difference in transmission from CYP across different settings, with the pooled SAR from CYP index cases in household studies (7.6%) being 10-fold higher than in school studies (0.7%), despite a similar quantity and quality of evidence in both settings. We were unable to draw conclusions about transmissibility from CYP compared with adults in educational settings, due to wide confidence intervals and lack of studies reporting adjusted analyses. We found no evidence that transmission differed from CYP index cases to contacts of differing ages. Similar to our findings, other studies have concluded that household settings have higher transmission potential than other settings such as schools.[17, 18] This disparity may reflect differences in the duration and intensity of social mixing within schools compared with households, with more prolonged, intense and intimate contacts between children and siblings or parents within households carrying a greater risk of transmission.[69] Our findings may also reflect the successful operation of NPI mitigations within schools in markedly reducing transmission.[70] This observation is supported by findings from some of the included school studies, including a lower prevalence in schools than in surrounding communities and the lack of notable clustering of infection within classrooms, even when local prevalence was high. Lack of clustering is supported by a number of studies not included in our review for quality reasons including a national study from Luxembourg.[71] There may, however, be systematic bias that might contribute to lower transmission in school compared with household studies. For example, CYP who are known to be infected or are contacts of positive cases are usually excluded from school but would be included within household studies. However, a substantial proportion of infected CYP are likely to be asymptomatic and, therefore, unlikely to be absent from school.[10] Biases related to relatively low numbers of CYP index cases, adequacy of contact-tracing and validity of PCR or serology testing in CYP apply equally to both school and household studies.

Our meta-regression findings that local community incidence was positively associated with school infection prevalence, as was incidence in the month prior, whereas seroprevalence was only associated with historical community incidence, show the inter-dependence of schools with their localities with respect to infection levels. Ismail et al.[72] reported the risk of an outbreak increased by 72% for every five cases per 100⍰000 population increase in community incidence, whilst Willeit et al.[56] reported that the odds of testing positive in schools were 1.64 (1.38, 1.96) for a two-fold higher community incidence. Our findings support the hypothesis that school infections predominantly reflect community infection levels, although our analysis could not attribute causality.

Our review included a number of studies undertaken when the prevalence of variants with higher transmissibility (e.g. alpha or B.1.1.7 variant) was rising or dominant, although most studies preceded this. No contact-tracing studies were included of transmission related to the delta variant although two school prevalence studies included data collection whilst delta infection was rising. Our findings therefore cannot be assumed to apply to periods when delta was predominant. However, whilst the delta variant has substantially higher overall transmissibility, and the prevalence of delta infection in children has been high at a time when adult populations had high vaccination coverage, there is no evidence of variant-specific differential transmission between children and adults. It is possible that the differential in transmission between school and household settings is lower for the higher transmissibility variants such as delta or omicron than reported here, although the higher transmissibility of the delta variant appears not to be setting-specific.

### Limitations

Our data are subject to a number of limitations. Potential biases in school studies have been discussed above. RT-PCR studies may under-estimate infection in children compared with serology,[36] and different seroassays may provide differing results. Many of the included studies, however, combined findings from both PCR and serology,[10, 31, 32, 39, 40, 44, 47, 48, 54, 67] or undertook repeated PCR measures[40, 44, 45, 49-51, 53, 60] Importantly, though, these issues are likely to be similar across both contact-tracing and population studies and, therefore, would not alter the notable differences we found by setting.

Contact-tracing studies are open to bias due to missed testing of contacts, although we only included those who planned routine testing of all contacts and who achieved a high proportion of contacts tested. Low numbers of child index cases and their contacts in some studies may also be a source of bias. Population studies may be biased by higher participation by higher socio-economic status groups and also as some studies specifically excluded those with recent contacts or symptoms.[50]

We conducted multi-level analyses accounting for the nesting of multiple rounds of data-collection within single studies. Some of the smaller meta-analyses, however, may have been overly influenced by studies with many rounds of testing. Meta-regression analyses are conducted at study rather than individual level and are, therefore, subject to ecological biases and cannot infer causality.

Our findings relate largely to the original/Wuhan virus and the alpha variant and it is unclear how generalisable they will be to the delta or other variants. Paucity of data meant we were unable to compare transmissibility from CYP between the Wuhan and alpha variants. Additionally all data precede widespread vaccination of adults and no studies included populations of teenagers who had been vaccinated. Our data were largely limited to high-income countries and there is an urgent need for similar studies from low-and-middle-income countries.

## Conclusions and implications

We found no difference in transmission of SARS-CoV-2 from CYP compared with adults within household settings. Secondary attack rates were markedly lower in school compared with household settings and there was little clustering of infections within schools, suggesting that household transmission is more high risk than school transmission in this pandemic.

School infection prevalence was associated with community infection incidence in the month before and during the study, with seroprevalence associated with historical community infections, supporting hypotheses that school infections broadly reflect community infections. These findings are important for guiding policy decisions on school operations during the pandemic. With appropriate mitigations, school infections can be limited and face-to-face learning is feasible, even at times of moderate to high community prevalence and in the presence of variants with higher transmissibility.

Our findings support a potential role for vaccination of CYP, if proven safe, in reducing transmission within households. Where countries go on to achieve very high levels of adult vaccination, this will focus transmission amongst the unvaccinated, increasing the relative importance of transmission amongst CYP.

Our findings largely relate to SARS-CoV-2 transmission from children before highly transmissible variants such as delta or omicron became predominant and this work needs replication once sufficient data are available from periods dominated by other variants. A number of other gaps in our knowledge remain about transmission from CYP, particularly relating to potential age-differences between younger and older children, and effectiveness of various NPIs, especially face masks, to reduce transmission in child-specific settings. Detailed population studies are required which link households and schools and use a combination of repeated PCR and serology testing to assess the risk of infection and direction of transmission across settings.

## Data Availability

All data are contained in the manuscript or available from the referenced published or preprint sources.

## Contributions

RV and CB conceptualised the paper, undertook the searches, contributed to data extraction and quality assessment, undertook the meta-analyses and led the writing of the manuscript. CW, OM, JC and JW contributed to eligibility assessment, data extraction and quality assessment. GMT and CB contributed to planning the analyses. All authors contributed to writing and editing of the manuscript.

## Declaration of interests

All authors declare no competing interests.

## Acknowledgements

We thank Kjetil Telle, Norwegian Institute of Public Health, and Marieke de Hoog, University Medical Center Utrecht, for providing additional data for their studies included here. We also thank Semina Michalopoulou and Zainab Dedat for checking the accuracy of data extraction.

## Notes

### Competing Interest Statement

The authors have declared no competing interest.

### Funding Statement

This study did not receive any funding

### Author Declarations

This study involves only data available from published or preprint sources

## References

1. Flasche S, Edmunds WJ. The role of schools and school-aged children in SARS-CoV-2 transmission. The Lancet Infectious diseases. 2021;21(3):298–9. Epub 2020/12/08. doi: 10.1016/S1473-3099(20)30927-0. PubMed PMID: 33306982.

2. Laxminarayan R, Wahl B, Dudala SR, Gopal K, Mohan C, Neelima S, et al. Epidemiology and transmission dynamics of COVID-19 in two Indian states. Science. 2020:eabd7672. doi: 10.1126/science.abd7672.

3. Okarska-Napierala M, Mandziuk J, Kuchar E. SARS-CoV-2 Cluster in Nursery, Poland. Emerg Infect Dis. 2021;27(1). Epub 2020/10/10. doi: 10.3201/eid2701.203849. PubMed PMID: 33035153; PubMed Central PMCID: PMCPMC7774538.

4. Fontanet A, Tondeur L, Madec Y, Grant R, Bescombes C, Jolly N, et al. Cluster of COVID-19 in northern France: A retrospective closed cohort study. medRxiv preprint server. 2020. Epub 23 April 2020. doi: 10.1101/2020.04.18.20071134.

5. Torres JP, Pinera C, De La Maza V, Lagomarcino AJ, Simian D, Torres B, et al. SARS-CoV-2 antibody prevalence in blood in a large school community subject to a Covid-19 outbreak: a cross-sectional study. Clin Infect Dis. 2020. Epub 2020/07/11. doi: 10.1093/cid/ciaa955. PubMed PMID: 32649743.

6. Stein-Zamir C, Abramson N, Shoob H, Libal E, Bitan M, Cardash T, et al. A large COVID-19 outbreak in a high school 10 days after schools’ reopening, Israel, May 2020. Euro Surveill. 2020;25(29). Epub 2020/07/29. doi: 10.2807/1560-7917.ES.2020.25.29.2001352. PubMed PMID: 32720636; PubMed Central PMCID: PMCPMC7384285.

7. Pray IW, Gibbons-Burgener SN, Rosenberg AZ, Cole D, Borenstein S, Bateman A, et al. COVID-19 Outbreak at an Overnight Summer School Retreat - Wisconsin, July-August 2020. MMWR Morb Mortal Wkly Rep. 2020;69(43):1600–4. Epub 2020/10/30. doi: 10.15585/mmwr.mm6943a4. PubMed PMID: 33119558; PubMed Central PMCID: PMCPMC7640998 Journal Editors form for disclosure of potential conflicts of interest. No other potential conflicts of interest were disclosed.

8. Szablewski CM, Chang KT, Brown MM, Chu VT, Yousaf AR, Anyalechi N, et al. SARS-CoV-2 Transmission and Infection Among Attendees of an Overnight Camp - Georgia, June 2020. MMWR Morb Mortal Wkly Rep. 2020;69(31):1023–5. Epub 2020/08/08. doi: 10.15585/mmwr.mm6931e1. PubMed PMID: 32759921; PubMed Central PMCID: PMCPMC7454898 Journal Editors form for disclosure of potential conflicts of interest. No potential conflicts of interest were disclosed.

9. Ulyte A, Radtke T, Abela IA, Haile SR, Berger C, Huber M, et al. Clustering and longitudinal change in SARS-CoV-2 seroprevalence in school children in the canton of Zurich, Switzerland: prospective cohort study of 55 schools. BMJ. 2021;372:m616. Epub 2021/03/19. doi: 10.1136/bmj.n616. PubMed PMID: 33731327; PubMed Central PMCID: PMCPMC7966948 at www.icmje.org/coi_disclosure.pdf and declare: support from Swiss School of Public Health (SSPH+), Swiss Federal Office of Public Health, private funders, funds of the cantons of Switzerland (Vaud, Zurich, and Basel), institutional funds of universities, and University of Zurich Foundation for the submitted work; no financial relationships with any organisations that might have an interest in the submitted work in the previous three years; no other relationships or activities that could appear to have influenced the submitted work.

10. Ladhani SN, Baawuah F, Beckmann J, Okike IO, Ahmad S, Garstang J, et al. SARS-CoV-2 infection and transmission in primary schools in England in June-December, 2020 (sKIDs): an active, prospective surveillance study. Lancet Child Adolesc Health. 2021. Epub 2021/03/20. doi: 10.1016/S2352-4642(21)00061-4. PubMed PMID: 33740430.

11. Viner RM, Mytton OT, Bonell C, Melendez-Torres GJ, Ward J, Hudson L, et al. Susceptibility to SARS-CoV-2 Infection Among Children and Adolescents Compared With Adults: A Systematic Review and Meta-analysis. JAMA pediatrics. 2020. Epub 2020/09/26. doi: 10.1001/jamapediatrics.2020.4573. PubMed PMID: 32975552.

12. Jarvis C, Munday J, Gimma A, Wong K, Van Zandvoort K, Funk S, et al. Social contacts in the UK from the CoMix social contact survey: Report for survey week 61. London School of Tropical Medicine and Hygiene, 2021 1 June 2021. Report No.

13. Suk JE, Vardavas C, Nikitara K, Phalkey R, Leonardi-Bee J, Pharris A, et al. The role of children in the transmission chain of SARS-CoV-2: a systematic review and update of current evidence. medRxiv. 2020:2020.11.06.20227264. doi: 10.1101/2020.11.06.20227264.

14. Gaythorpe K, Bhatia S, Mangal T, al. e. Children’s role in the COVID-19 pandemic: as systematic review of early surveillance data on susceptibility, severity, and transmissibility. London: Imperial College London, 2020 19-11-2020. Report No.

15. Xu W, Li X, Dozier M, He Y, Kirolos A, Lang Z, et al. What is the evidence for transmission of COVID-19 by children in schools? A living systematic review. medRxiv. 2020:2020.10.11.20210658. doi: 10.1101/2020.10.11.20210658.

16. Spielberger BD, Goerne T, Geweniger A, Henneke P, Elling R. Intra-Household and Close-Contact SARS-CoV-2 Transmission Among Children - a Systematic Review. Front Pediatr. 2021;9:613292. Epub 2021/04/27. doi: 10.3389/fped.2021.613292. PubMed PMID: 33898355; PubMed Central PMCID: PMCPMC8062727.

17. Madewell ZJ, Yang Y, Longini IM, Jr., Halloran ME, Dean NE. Household Transmission of SARS-CoV-2: A Systematic Review and Meta-analysis. JAMA network open. 2020;3(12):e2031756–e. doi: 10.1001/jamanetworkopen.2020.31756. PubMed PMID: 33315116.

18. Thompson HA, Mousa A, Dighe A, Fu H, Arnedo-Pena A, Barrett P, et al. SARS-CoV-2 setting-specific transmission rates: a systematic review and meta-analysis. Clinical infectious diseases : an official publication of the Infectious Diseases Society of America. 2021:ciab100. doi: 10.1093/cid/ciab100. PubMed PMID: 33560412.

19. Krishnaratne S, Pfadenhauer LM, Coenen M, Geffert K, Jung-Sievers C, Klinger C, et al. Measures implemented in the school setting to contain the COVID-19 pandemic: a rapid scoping review. Cochrane Database of Systematic Reviews. 2020;(12). doi: 10.1002/14651858.CD013812. PubMed PMID: CD013812.

20. Stuart EA, Dowdy DW. Evidence-based COVID-19 policy-making in schools. Nature Medicine. 2021. doi: 10.1038/s41591-021-01585-2.

21. Haag L, Blankenburg J, Unrath M, Grabietz J, Kahre E, Galow L, et al. Prevalence and Transmission of SARS-CoV-2 in Childcare Facilities: A Longitudinal Study. medRxiv. 2021:2021.04.16.21255616. doi: 10.1101/2021.04.16.21255616.

22. Walsh S, Chowdhury A, Russell S, Braithwaite V, Ward J, Waddington C, et al. Do school closures reduce community transmission of COVID-19? A systematic review of observational studies. medRxiv. 2021:2021.01.02.21249146. doi: 10.1101/2021.01.02.21249146.

23. Checklist for prevalence studies: The Joanna Briggs Institute Critical Appraisal tools for use in JBI Systematic Reviews. Adelaide, South Australia: Joanna Briggs Institute, 2017.

24. Munn Z, Moola S, Lisy K, Riitano D, Tufanaru C. Methodological guidance for systematic reviews of observational epidemiological studies reporting prevalence and cumulative incidence data. Int J Evid Based Healthc. 2015;13(3):147–53. Epub 2015/09/01. doi: 10.1097/XEB.0000000000000054. PubMed PMID: 26317388.

25. Loney PL, Chambers LW, Bennett KJ, Roberts JG, Stratford PW. Critical appraisal of the health research literature: prevalence or incidence of a health problem. Chronic Dis Can. 1998;19(4):170–6. Epub 1999/02/24. PubMed PMID: 10029513.

26. Moola S, Munn Z, Tufanaru C, Aromataris E, Sears K, Sfetcu R, et al. Systematic reviews of etiology and risk. In: Aromataris E, Munn Z, editors. JBI Manual for Evidence Synthesis: JBI; 2020.

27. Blaisdell LL, Cohn W, Pavell JR, Rubin DS, Vergales JE. Preventing and Mitigating SARS-CoV-2 Transmission - Four Overnight Camps, Maine, June-August 2020. MMWR Morbidity and mortality weekly report. 2020;69(35):1216–20. doi: 10.15585/mmwr.mm6935e1. PubMed PMID: 32881850.

28. Schoeps A, Hoffmann D, Tamm C, Vollmer B, Haag S, Kaffenberger T, et al. COVID-19 transmission in educational institutions August to December 2020 in Germany: a study of index cases and close contact cohorts. medRxiv. 2021:2021.02.04.21250670. doi: 10.1101/2021.02.04.21250670.

29. Yoon Y, Kim KR, Park H, Kim S, Kim YJ. Stepwise School Opening and an Impact on the Epidemiology of COVID-19 in the Children. Journal of Korean medical science. 2020;35(46):e414. Epub 2020/12/02. doi: 10.3346/jkms.2020.35.e414. PubMed PMID: 33258334; PubMed Central PMCID: PMCPMC7707922.

30. Larosa E, Djuric O, Cassinadri M, Cilloni S, Bisaccia E, Vicentini M, et al. Secondary transmission of COVID-19 in preschool and school settings in northern Italy after their reopening in September 2020: a population-based study. Euro Surveill. 2020;25(49). doi: 10.2807/1560-7917.ES.2020.25.49.2001911. PubMed PMID: 33303065; PubMed Central PMCID: PMCPMC7730487.

31. National Centre for Immunisation Research and Surveillance or NCIRS. COVID-19 in schools and early childhood education and care services – the Term 3 experience in NSW. Sydney, Australia: National Centre for Immunisation Research and Surveillance, 2020 21 October 2020. Report No.

32. National Centre for Immunisation Research and Surveillance or NCIRS. COVID-19 in schools and early childhood education and care services – the Term 4 experience in NSW. Sydney, Australia: National Centre for Immunisation Research and Surveillance, 2021 9 March 2021. Report No.

33. Brandal LT, Ofitserova TS, Meijerink H, Rykkvin R, Lund HM, Hungnes O, et al. Minimal transmission of SARS-CoV-2 from paediatric COVID-19 cases in primary schools, Norway, August to November 2020. Euro surveillance : bulletin Europeen sur les maladies transmissibles = European communicable disease bulletin. 2021;26(1):2002011. doi: 10.2807/1560-7917.ES.2020.26.1.2002011. PubMed PMID: 33413743.

34. Park YJ, Choe YJ, Park O, Park SY, Kim YM, Kim J, et al. Contact Tracing during Coronavirus Disease Outbreak, South Korea, 2020. Emerg Infect Dis. 2020;26(10). Epub 2020/07/17. doi: 10.3201/eid2610.201315. PubMed PMID: 32673193.

35. Hu S, Wang W, Wang Y, Litvinova M, Luo K, Ren L, et al. Infectivity, susceptibility, and risk factors associated with SARS-CoV-2 transmission under intensive contact tracing in Hunan, China. Nat Commun. 2021;12(1):1533. Epub 2021/03/23. doi: 10.1038/s41467-021-21710-6. PubMed PMID: 33750783; PubMed Central PMCID: PMCPMC7943579 Seqirus, and H.Y. has received research funding from Sanofi Pasteur, GlaxoSmithKline, Yichang HEC Changjiang Pharmaceutical Company, and Shanghai Roche Pharmaceutical Company. None of those research funding is related to COVID-19. All other authors report no competing interests.

36. Dattner I, Goldberg Y, Katriel G, Yaari R, Gal N, Miron Y, et al. The role of children in the spread of COVID-19: Using household data from Bnei Brak, Israel, to estimate the relative susceptibility and infectivity of children. PLoS computational biology. 2021;17(2):e1008559–e. doi: 10.1371/journal.pcbi.1008559. PubMed PMID: 33571188.

37. Li F, Li Y-Y, Liu M-J, Fang L-Q, Dean NE, Wong GWK, et al. Household transmission of SARS-CoV-2 and risk factors for susceptibility and infectivity in Wuhan: a retrospective observational study. Lancet Infect Dis. 2021. doi: 10.1016/S1473-3099(20)30981-6.

38. Kim J, Choe YJ, Lee J, Park YJ, Park O, Han MS, et al. Role of children in household transmission of COVID-19. Arch Dis Child. 2020. Epub 2020/08/10. doi: 10.1136/archdischild-2020-319910. PubMed PMID: 32769089.

39. Verberk J, de Hoog M, Westerhof I, Van Goethem S, Lammens C, Ieven M, et al. Transmission of SARS-CoV-2 within households: a prospective cohort study in the Netherlands and Belgium – Interim results. medRxiv. 2021:2021.04.23.21255846. doi: 10.1101/2021.04.23.21255846.

40. Reukers DFM, van Boven M, Meijer A, Rots N, Reusken C, Roof I, et al. High infection secondary attack rates of SARS-CoV-2 in Dutch households revealed by dense sampling. Clin Infect Dis. 2021. Epub 2021/04/07. doi: 10.1093/cid/ciab237. PubMed PMID: 33822007; PubMed Central PMCID: PMCPMC8083540.

41. Lyngse FP, Mølbak K, Træholt Frank K, Nielsen C, Skov RL, Kirkeby CT. Association between SARS-CoV-2 Transmission Risk, Viral Load, and Age: A Nationwide Study in Danish Households. medRxiv. 2021:2021.02.28.21252608. doi: 10.1101/2021.02.28.21252608.

42. Telle K, Jørgensen SB, Hart R, Greve-Isdahl M, Kacelnik O. Secondary attack rates of COVID-19 in Norwegian families: a nation-wide register-based study. European journal of epidemiology. 2021:1–8. doi: 10.1007/s10654-021-00760-6. PubMed PMID: 34036466.

43. Varma JK, Thamkittikasem J, Whittemore K, Alexander M, Stephens DH, Arslanian K, et al. COVID-19 Infections among Students and Staff in New York City Public Schools. Pediatrics. 2021:e2021050605. doi: 10.1542/peds.2021-050605.

44. Jordan I, de Sevilla MF, Fumado V, Bassat Q, Bonet-Carne E, Fortuny C, et al. Transmission of SARS-CoV-2 infection among children in summer schools applying stringent control measures in Barcelona, Spain. Clinical Infectious Diseases. 2021. doi: 10.1093/cid/ciab227.

45. Hoehl S, Kreutzer E, Schenk B, Westhaus S, Foppa I, Herrmann E, et al. Longitudinal testing for respiratory and gastrointestinal shedding of SARS-CoV-2 in day care centres in Hesse, Germany. Clin Infect Dis. 2021. Epub 2021/01/04. doi: 10.1093/cid/ciaa1912. PubMed PMID: 33388748; PubMed Central PMCID: PMCPMC7799213.

46. Kriemler S, Ulyte A, Ammann P, Peralta GP, Berger C, Puhan MA, et al. Surveillance of Acute SARS-CoV-2 Infections in School Children and Point-Prevalence During a Time of High Community Transmission in Switzerland. Front Pediatr. 2021;9:645577. Epub 2021/04/03. doi: 10.3389/fped.2021.645577. PubMed PMID: 33796490; PubMed Central PMCID: PMCPMC8007924.

47. Theuring S, Thielecke M, van Loon W, Hommes F, Hülso C, von der Haar A, et al. SARS-CoV-2 infection and transmission in school settings during the second wave in Berlin, Germany: a cross-sectional study. medRxiv. 2021:2021.01.27.21250517. doi: 10.1101/2021.01.27.21250517.

48. Thielecke M, Theuring S, van Loon W, Hommes F, Mall MA, Rosen A, et al. SARS-CoV-2 infections in kindergartens and associated households at the start of the second wave in Berlin, Germany - a cross sectional study. European journal of public health. 2021:ckab079. doi: 10.1093/eurpub/ckab079. PubMed PMID: 33956945.

49. Hoch M, Vogel S, Kolberg L, Dick E, Fingerle V, Eberle U, et al. Weekly SARS-CoV-2 Sentinel Surveillance in Primary Schools, Kindergartens, and Nurseries, Germany, June⍰November 2020. Emerg Infect Dis. 2021;27(8). Epub 2021/06/05. doi: 10.3201/eid2708.204859. PubMed PMID: 34087088.

50. Lübke N, Schupp A-K, Bredahl R, Kraus U, Hauka S, Andrée M, et al. Screening for SARS-CoV-2 infections in daycare facilities for children in a large city in Germany. medRxiv. 2021:2021.02.26.21252510. doi: 10.1101/2021.02.26.21252510.

51. Doron S, Ingalls RR, Beauchamp A, Boehm J, Boucher HW, Chow LH, et al. Weekly SARS-CoV-2 screening of asymptomatic students and staff to guide and evaluate strategies for safer in-person learning. medRxiv. 2021:2021.03.20.21253976. doi: 10.1101/2021.03.20.21253976.

52. COVID-19 Schools Infection Survey Round 1, England: November 2020. England: Office for National Statistics (ONS), 2020 17 Dec 2020. Report No.

53. Villani A, Coltella L, Ranno S, Bianchi di Castelbianco F, Murru PM, Sonnino R, et al. School in Italy: a safe place for children and adolescents. Ital J Pediatr. 2021;47(1):23. Epub 2021/02/04. doi: 10.1186/s13052-021-00978-w. PubMed PMID: 33531046; PubMed Central PMCID: PMCPMC7851807.

54. Hommes F, van Loon W, Thielecke M, Abramovich I, Lieber S, Hammerich R, et al. SARS-CoV-2 Infection, Risk Perception, Behaviour and Preventive Measures at Schools in Berlin, Germany, during the Early Post-Lockdown Phase: A Cross-Sectional Study. Int J Environ Res Public Health. 2021;18(5). Epub 2021/04/04. doi: 10.3390/ijerph18052739. PubMed PMID: 33800392; PubMed Central PMCID: PMCPMC7967466.

55. Kirsten C, Unrath M, Lück C, Dalpke AH, Berner R, Armann J. SARS-CoV-2 seroprevalence in students and teachers: a longitudinal study from May to October 2020 in German secondary schools. BMJ open. 2021;11(6):e049876. Epub 2021/06/12. doi: 10.1136/bmjopen-2021-049876. PubMed PMID: 34112645; PubMed Central PMCID: PMCPMC8193693.

56. Willeit P, Krause R, Lamprecht B, Berghold A, Hanson B, Stelzl E, et al. Prevalence of RT-qPCR-detected SARS-CoV-2 infection at schools: First results from the Austrian School-SARS-CoV-2 prospective cohort study. The Lancet Regional Health - Europe. 2021;5:100086. doi: https://doi.org/10.1016/j.lanepe.2021.100086.

57. Fontanet A, Tondeur L, Grant R, Temmam S, Madec Y, Bigot T, et al. SARS-CoV-2 infection in schools in a northern French city: a retrospective serological cohort study in an area of high transmission, France, January to April 2020. Euro Surveill. 2021;26(15). Epub 2021/04/17. doi: 10.2807/1560-7917.ES.2021.26.15.2001695. PubMed PMID: 33860747; PubMed Central PMCID: PMCPMC8167414.

58. Lachassinne E, de Pontual L, Caseris M, Lorrot M, Guilluy C, Naud A, et al. SARS-CoV-2 transmission among children and staff in daycare centres during a nationwide lockdown in France: a cross-sectional, multicentre, seroprevalence study. The Lancet Child & Adolescent Health. 2021;5(4):256–64. doi: 10.1016/S2352-4642(21)00024-9.

59. Ladhani SN, Ireland G, Baawuah F, Beckmann J, Okike IO, Ahmad S, et al. Emergence of SARS-CoV-2 Alpha (B.1.1.7) variant, infection rates, antibody seroconversion and seroprevalence rates in secondary school students and staff: active prospective surveillance, December 2020 to March 2021, England. medRxiv. 2021:2021.07.14.21260496. doi: 10.1101/2021.07.14.21260496.

60. House T, Pellis L, Pouwels KB, Bacon S, Eidukas A, Jahanshahi K, et al. Inferring Risks of Coronavirus Transmission from Community Household Data2021 April 01, 2021:[2104.04605 p.]. Available from: https://ui.adsabs.harvard.edu/abs/2021arXiv210404605H.

61. Espenhain L, Tribler S, Jørgensen CS, Holm Hansen C, Wolff Sönksen U, Ethelberg S. Prevalence of SARS-CoV-2 antibodies in Denmark 2020: results from nationwide, population-based sero-epidemiological surveys. medRxiv. 2021:2021.04.07.21254703. doi: 10.1101/2021.04.07.21254703.

62. Yoon Y, Kim K-R, Park H, Kim Sy, Kim Y-J. Stepwise School Opening Online and Off-line and an Impact on the Epidemiology of COVID-19 in the Pediatric Population. medRxiv. 2020:2020.08.03.20165589. doi: 10.1101/2020.08.03.20165589.

63. Telle K, Jørgensen SB, Hart R, Greve-Isdahl M, Kacelnik O. Secondary attack rates of COVID-19 in Norwegian families: A nation-wide register-based study. medRxiv. 2021:2021.03.06.21252832. doi: 10.1101/2021.03.06.21252832.

64. COVID-19 Infection Survey: methods and further information. England: Office for National Statistics (ONS), 2021 26 March 2021. Report No.

65. Ulyte A, Radtke T, Abela IA, Haile SR, Ammann P, Berger C, et al. Evolution of SARS-CoV-2 seroprevalence and clusters in school children from June 2020 to April 2021 reflect community transmission: prospective cohort study <em>Ciao Corona</em>. medRxiv. 2021:2021.07.19.21260644. doi: 10.1101/2021.07.19.21260644.

66. Thielecke M, Theuring S, van Loon W, Hommes F, Mall MA, Rosen A, et al. SARS-CoV-2 infections in kindergartens and associated households at the start of the second wave in Berlin, Germany – a cross sectional study. medRxiv. 2020:2020.12.08.20245910. doi: 10.1101/2020.12.08.20245910.

67. COVID-19 Schools Infection Survey Rounds 2 to 5. England: Office for National Statistics (ONS), 2021 July 2021. Report No.

68. Fontanet A, Grant R, Tondeur L, Madec Y, Grzelak L, Cailleau I, et al. SARS-CoV-2 infection in primary schools in northern France: A retrospective cohort study in an area of high transmission. medRxiv. 2020:2020.06.25.20140178. doi: 10.1101/2020.06.25.20140178.

69. Mossong J, Hens N, Jit M, Beutels P, Auranen K, Mikolajczyk R, et al. Social contacts and mixing patterns relevant to the spread of infectious diseases. PLoS Med. 2008;5(3):e74. Epub 2008/03/28. doi: 10.1371/journal.pmed.0050074. PubMed PMID: 18366252; PubMed Central PMCID: PMCPMC2270306.

70. Lessler J, Grabowski MK, Grantz KH, Badillo-Goicoechea E, Metcalf CJE, Lupton-Smith C, et al. Household COVID-19 risk and in-person schooling. Science. 2021;372(6546):1092–7. Epub 2021/05/01. doi: 10.1126/science.abh2939. PubMed PMID: 33927057; PubMed Central PMCID: PMCPMC8168618.

71. Mossong J, Mombaerts L, Veiber L, Pastore J, Coroller GL, Schnell M, et al. SARS-CoV-2 transmission in educational settings during an early summer epidemic wave in Luxembourg, 2020. BMC Infect Dis. 2021;21(1):417. Epub 2021/05/06. doi: 10.1186/s12879-021-06089-5. PubMed PMID: 33947340; PubMed Central PMCID: PMCPMC8093902.

72. Ismail SA, Saliba V, Lopez Bernal J, Ramsay ME, Ladhani SN. SARS-CoV-2 infection and transmission in educational settings: a prospective, cross-sectional analysis of infection clusters and outbreaks in England. The Lancet Infectious diseases. 2021;21(3):344–53. Epub 2020/12/08. doi: 10.1016/S1473-3099(20)30882-3. PubMed PMID: 33306981.

