## Supplement for "Transmission of SARS-CoV-2 by children and young people in households and schools: a meta-analysis of population-based and contact-tracing studies"

### Appendix

#### Table 1. Search terms

1. PubMed: ("COVID-19"[Text Word] OR "2019-nCoV"[Text Word] OR "SARS-CoV-2"[Text Word]) AND ("child*"[All Fields] OR "infant*"[All Fields]) AND ("disease transmission, infectious"[MeSH Terms] OR "epidemiology"[MeSH Terms] OR "schools"[MeSH Terms])

2. medRxiv: separate searches were undertaken for ‘COVID-19 & CHILD & transmission’ , ‘COVID-19 & CHILD & epidemiology’, ‘COVID-19 & School’ using ‘all words’ in title, abstract or full-text

3. the COVID-19 Living Evidence database (<https://zika.ispm.unibe.ch/assets/data/pub/search_beta/>) using the terms ‘school AND child’

4. Europe PMC (preprints): (("2019-nCoV" OR "2019nCoV" OR "COVID-19" OR "SARS-CoV-2" OR "COVID19" OR "COVID" OR "SARS-nCoV") AND "school" AND "Child") AND (SRC:PPR)

#### Table 2. Data notes and source of data for each study

Population studies

| ***Study*** | ***Data notes*** |
| --- | --- |
| Villani | Table 1 & text. Total N of 1083 used for all rounds. |
| Ulyte | Published paper Table 2 for R1 & 2. Note some differing figures given across the preprints and published paper and some differences with retrospective reporting of R1 & 2 from the R3 preprint. R3: data from preprint Appendix 2 |
| Kriemler | Table 1 & text |
| Hommes | Data from text. Note that N =382 as 3 students refused both PCR and serology |
| Theuring | Table 1 |
| Kirsten | Table 1 |
| Thielecke | Text. Note N=152 as swabs only taken for 152/155. Total N for sample (155) used for serology as not otherwise stated |
| Hoch R1 & 2 | Data from Figure 2 & text. N calculated for R1 & 2 from Figure 2. Both rounds together = 2149 swabs. |
| Willeit | Table 2 & Section 3.2 |
| Varma | Appendix eTable 2 |
| Schools Infection Survey (SIS) | R1: Table 1a. Note the repeat cross-sectional data for each round were used. There is some variation in retrospective reporting of findings from previous rounds when a new round is published as data were reported including onlly local authorities that took part in the later rounds. For all SIS data, prevalence was reported therefore the number of positive children was calculated. R1 serology reported in R2 report and online spreadsheet Table 2a.  R2: Table 1a of online spreadsheet. R2 seroprevalence reported in R5 online spreadsheet Table 2b  R4: Table 1a of online spreadsheet  R5: Table 1a of online spreadsheet. |
| Ladhani sKIDs | PCR data from Table 1 & p4; seroloprevalence p5 |
| Fontanet | Table 1 |
| Jordan | Active surveillance data only used for population prevalence data. Results section first paragraph |
| Lubke | Data p5&6. Note used N =3765 that gave samples rather than N that participated. |
| Hoehl | R1: Table 1 & Results text  R2 & 3: data in tet |
| Doron | Table 1. Note we included both students positive by screening and those positive from outside testing. The N for weeks 6-18 (Rounds 2 & 3) is stated as 2403 in Table 1. |
| Ladhani_sKIDsPLUS | Serology data for R1-3 in Figure 1 (figure table). Note Abbott assay used across R1-3 for consistency  R3: PCR Data in Results text |
| Lachassine | Data in Results text |

Contact tracing studies

| **Study** | **Data notes** |
| --- | --- |
| Reukers | Age-dependent transmissibility model data taken from Table 3. |
| Hu | Table 2 |
| Dattner | Table 1 plus Section 3.4 |
| Kim | Results first paragraph. Note 40 contacts with same exposure as index removed from analysis. |
| Laxminarayan | Only data from index cases with tested contacts used - from Table S3. Data from both Tamil Nadu and Andhra Pradesh. |
| Lyngse | Table 1. |
| Park | Tables 1 & 2. Household contacts only were included. Adult comparator 20-59y |
| Li | Table 2. Used data from households with a single primary case. In our unadjusted meta-analysis, we used a comparison group of all adults 20 plus in order to be more comparable with other studies in the analysis; whereas Li used >=60 years as reference group in their adjusted chain-binomial models (data Table S13) |
| Telle | Table 1 and Supplement Table A. SAR14 used for comparison with other studies. Only 0-16yo index cases included due to low testing of contacts amongst 17-20 year olds. Adult data were taken from the Parent category. |
| Brandal | Data from text p2. |
| Schoeps | Table 1. Data used for 441 index cases with contact data |
| Larosa | Table 2. Data used for ‘all students’ and ‘teachers/staff’ |
| Yoon | Table 5. |
| Jordan | Data from Table 1 & 2. Data from Recruitment Pathway 1 (RP1) only used |
| Macartney | Data extracted from Figures 3-6 of both Term 3 and Term 4 reports. Data summarized as early years plus primary (=Child) and secondary school |
| Blaisdell | Result text p.1218. Index cases identified after arrival at camp. Their cohort bubbles defined as their contacts. |
| Verberk | Child data used from data supplied by authors, which contained additional cases to those in the preprint. Adult data not supplied. Household contacts only included. Missing data on household size for one child index was replaced with median household size across 38 other index child cases. |
| House | Data from Tranche 3 and 4 only used. Data from Table 3 and Figure 6 |
| Lyngse | Table 2. OR recalculated to use 30-34y as adult reference category to match previous version of this analysis (as June 2021 update used 15-20 year olds, an inappropriate reference category for our analyses). Sample sizes for this from Appendix Table S2. |

#### Quality assessment

Methodological quality was independently assessed by two authors (RV and CW) using a score adapted from previously published quality assessment (See Appendix Table 2). Disagreement was resolved by consensus. The score was based on quality of study design and methodology, particularly with regard to the adequacy of study population in meeting the study aims and participant recruitment and testing; and assessment of risk of bias and generalisability of the study results. Key potential biases in the context of this study were identified as being 1) the risk of under-ascertainment of cases of infection due to asymptomatic or minimally symptomatic infection; and 2) representativeness of the study sample, with low rates of testing and/or small sample size and/or skewed study populations all leading to potentially biased results. Studies were categorised as high quality if they scored >=10/12; medium if they scored >=7-9; and low if they scored <7 (see Appendix Table 3).

#### Appendix Table 3. Quality scoring criteria

A. Quality scoring for population studies:

|  | Yes (1 point) | No or unclear (0 points) | Not applicable |
| --- | --- | --- | --- |
| Is the study aim/objective stated clearly in the abstract, introduction, or methods section? |  |  |  |
| Was the target study population (sampling frame) suitable for the objectives of the study, and were the inclusion and exclusion criteria clearly described and appropriate? |  |  |  |
| Was the planned sample size adequate? |  |  |  |
| Was the study setting, including school-based mitigation strategies to prevent infection transmission (where relevant) described in adequate detail? |  |  |  |
| Were the methods of testing appropriate (RT-PCR or validated serology)? |  |  |  |
| Were asymptomatic and minimally symptomatic infections as likely to be detected as symptomatic infections (i.e. teating strategy not based on presence of symptoms)? |  |  |  |
| Were the statistical tests used to assess the relevant outcomes appropriate? |  |  |  |
| Are the participant characteristics described in suitable detail? (e.g. number of participants, age, and gender, relevant subgroups) |  |  |  |
|  | Highly representative (2 points) e.g. >=75% of total sample population enrolled and tested OR large randomly selected or stratified sample of source population enrolled and tested | Somewhat representative (1 point)  e.g. 40-75% of planned or eligible population enrolled and tested | Poorly representative or unclear (0 points)  <40% of potential enrolled and/or tested or unclear |
| How representative was the recruited/ tested population of the underlying source population? |  |  |  |
|  | High (2 points) – large, nationally representative sample; | Medium (score 1 point)- medium sized and/or regionally representative sample | Low (score 0) - small and/or selective group |
| How generalisable are results from the study participants to the wider population? |  |  |  |
| Total score: |  | | |

B. **Contact tracing studies:**

|  | Yes | No or unclear | Not applicable |
| --- | --- | --- | --- |
| Is the study aim/objective stated clearly in the abstract, introduction, or methods section? |  |  |  |
| Were the index cases identified suitable for the objectives of the study? Was the index case definition clear and appropriate? |  |  |  |
| Were contacts of cases appropriately defined, identified and tested at an appropriate time point regardless of symptoms? |  |  |  |
| Was the sample size adequate? |  |  |  |
| Was the study setting from described in relevant detail? Were case isolation and school-based mitigation strategies described in adequate detail? |  |  |  |
| Were the methods of testing appropriate (RT-PCR or validated serology)? |  |  |  |
| Were the statistical tests used to assess the relevant outcomes appropriate? |  |  |  |
| Are the study participants characteristics described in suitable detail? (e.g. number of participants, age, and gender, relevant subgroups) |  |  |  |
|  | Highly representative (2 points) e.g. >=85% of contacts enrolled and tested OR large randomly selected or stratified sample of contacts enrolled and tested | Somewhat representative (1 point)  e.g. >=50-85% of contacts enrolled and tested | Poorly representative or unclear (0 points)  <50% of contactsenrolled and/or tested or unclear |
| Were representative was the recruited/ tested contacts of the target study population? |  |  |  |
|  | High (2 points) – large, nationally representative sample; | Medium (score 1 point)- medium sized and/or regionally representative sample | Low (score 0) - small and/or selective group |
| How generalisable are results from the study participants to the wider population? |  |  |  |
| Total score |  | | |

#### Appendix Table 4. Quality scores of included studies

Note that quality ratings may refer to subsets of data used in our analyses, e.g. where a paper reports both higher quality and lower quality data (in terms of contact tracing or testing), our quality rating refers to the data we include in these analyses rather than the whole paper.

A. Contact-tracing studies

|  | Is the study aim/objective stated clearly in the abstract, introduction, or methods section? | Were the index cases identified suitable for the objectives of the study? Was the index case definition clear and appropriate? | Were contacts of cases appropriately defined, identified and tested at an appropriate time point regardless of symptoms? | Was the sample size adequate? | Was the study setting from described in relevant detail? Were case isolation and school-based mitigation strategies described in adequate detail? | Were the methods of testing appropriate (RT-PCR or validated serology)? | Were the statistical tests used to assess the relevant outcomes appropriate? | Are the study participants characteristics described in suitable detail? (e.g. number of participants, age, and gender, relevant subgroups) | Were representative was the recruited/ tested contacts of the target study population? | How generalisable are results from the study participants to the wider population? | Score | Quality | **Notes** |
| --- | --- | --- | --- | --- | --- | --- | --- | --- | --- | --- | --- | --- | --- |
| Brandal | 1 | 1 | 1 | 1 | 0 | 1 | 1 | 1 | 1 | 1 | 9 | Medium |  |
| Schoeps | 1 | 1 | 1 | 1 | 1 | 1 | 1 | 1 | 2 | 1 | 11 | High | Quality assessed on subset of cases (n=441) with information on all contacts. |
| Hu | 1 | 1 | 1 | 1 | 1 | 1 | 1 | 1 | 0 | 1 | 9 | Medium |  |
| Reukers | 1 | 1 | 1 | 1 | 1 | 1 | 1 | 0 | 1 | 1 | 9 | Medium |  |
| Larosa | 1 | 1 | 0 | 1 | 1 | 1 | 0 | 0 | 2 | 1 | 8 | Medium |  |
| Lyngse | 1 | 1 | 0 | 1 | 1 | 1 | 1 | 1 | 1 | 2 | 10 | High |  |
| Macartney (T3, T4) | 1 | 1 | 1 | 1 | 1 | 1 | 1 | 0 | 2 | 1 | 10 | High |  |
| Park | 1 | 1 | 1 | 1 | 1 | 1 | 1 | 0 | 2 | 2 | 11 | High |  |
| Li | 1 | 1 | 0 | 1 | 1 | 1 | 1 | 0 | 2 | 1 | 9 | Medium | Household contacts were routinely tested for the majority but not all of the study period, thus Risk of bias 1 rated as Uncertain however the study was included |
| Telle | 1 | 1 | 1 | 1 | 1 | 1 | 1 | 1 | 2 | 2 | 12 | High | Quality rating refers to contacts of child index cases which had high (>85%) proportions of contacts tested. |
| Dattner | 1 | 1 | 1 | 1 | 1 | 1 | 1 | 1 | 2 | 1 | 11 | High |  |
| Kim | 1 | 1 | 1 | 1 | 1 | 1 | 1 | 1 | 2 | 2 | 12 | High |  |
| Laxminarayan | 1 | 0 | 1 | 1 | 1 | 1 | 1 | 1 | 1 | 1 | 9 | Medium |  |
| Yoon | 1 | 1 | 1 | 1 | 1 | 1 | 1 | 1 | 2 | 2 | 12 | High |  |
| Blaisdell | 1 | 1 | 1 | 1 | 1 | 1 | 1 | 1 | 2 | 1 | 11 | High |  |
| Verberk | 1 | 1 | 1 | 1 | 1 | 1 | 1 | 1 | 2 | 1 | 11 | High |  |
| Varma et al. | 1 | 1 | 1 | 1 | 1 | 1 | 1 | 0 | 0 | 1 | 8 | Medium |  |
| Jordan | 1 | 1 | 1 | 1 | 1 | 1 | 1 | 1 | 2 | 1 | 11 | High |  |

Population prevalence studies

|  | Is the study aim/objective stated clearly in the abstract, introduction, or methods section? | Was the target study population (sampling frame) suitable for the objectives of the study, and were the inclusion and exclusion criteria clearly described and appropriate? | Was the planned sample size adequate? | Was the study setting, including school-based mitigation strategies to prevent infection transmission (where relevant) described in adequate detail? | Were the methods of testing appropriate (RT-PCR or validated serology)? | Were asymptomatic and minimally symptomatic infections as likely to be detected as symptomatic infections (i.e. teating strategy not based on presence of symptoms)? | Were the statistical tests used to assess the relevant outcomes appropriate? | Are the participant characteristics described in suitable detail? (e.g. number of participants, age, and gender, relevant subgroups) | How representative was the recruited/ tested population of the underlying source population? | How generalisable are results from the study participants to the wider population? | Score |  |
| --- | --- | --- | --- | --- | --- | --- | --- | --- | --- | --- | --- | --- |
| Villani | 1 | 1 | 1 | 0 | 1 | 1 | 1 | 0 | 2 | 0 | 8 | Medium |
| Kriemler | 1 | 1 | 1 | 1 | 1 | 1 | 1 | 1 | 1 | 1 | 10 | High |
| Uylte | 1 | 1 | 1 | 1 | 1 | 1 | 1 | 1 | 1 | 1 | 10 | High |
| Hommes | 1 | 1 | 1 | 1 | 1 | 1 | 1 | 1 | 2 | 1 | 11 | High |
| Theuring | 1 | 1 | 1 | 1 | 1 | 1 | 1 | 1 | 1 | 1 | 10 | High |
| Armann A | 1 | 1 | 1 | 0 | 1 | 1 | 1 | 1 | 0 | 1 | 8 | Medium |
| Thielecke | 1 | 1 | 1 | 1 | 1 | 1 | 1 | 1 | 1 | 1 | 10 | High |
| Hoch | 1 | 1 | 1 | 1 | 1 | 1 | 1 | 1 | 1 | 1 | 10 | High |
| Willeit | 1 | 1 | 1 | 1 | 1 | 1 | 1 | 1 | 1 | 1 | 10 | High |
| CIS ONS | 1 | 1 | 1 | 1 | 1 | 1 | 1 | 1 | 1 | 2 | 11 | High |
| Ladhani sKIDs | 1 | 0 | 1 | 1 | 1 | 1 | 1 | 0 | 1 | 1 | 8 | Medium |
| House | 1 | 1 | 1 | 1 | 1 | 1 | 1 | 1 | 1 | 2 | 11 | High |
| Lubke | 1 | 1 | 1 | 1 | 1 | 1 | 1 | 1 | 2 | 1 | 11 | High |
| Hoehl | 1 | 1 | 1 | 1 | 0 | 1 | 1 | 1 | 1 | 1 | 9 | Medium |
| Espenhain | 1 | 1 | 1 | 1 | 1 | 1 | 1 | 1 | 2 | 1 | 11 | High |
| Doron | 1 | 1 | 1 | 1 | 1 | 1 | 1 | 1 | 1 | 1 | 10 | High |
| Fontanet | 1 | 1 | 1 | 1 | 1 | 1 | 1 | 1 | 1 | 1 | 10 | High |
| Varma | 1 | 1 | 1 | 1 | 1 | 1 | 1 | 0 | 1 | 1 | 9 | Medium |
| Jordan | 1 | 1 | 1 | 1 | 1 | 1 | 1 | 1 | 1 | 1 | 10 | High |
| Ladhani sKIDsPLUS | 1 | 0 | 1 | 1 | 1 | 1 | 1 | 0 | 1 | 1 | 8 | Medium |
| Lachassine | 1 | 1 | 1 | 1 | 1 | 1 | 1 | 1 | 0 | 1 | 9 | Medium |

#### Appendix Table 5. Community infection data and sources for school-based studies

The primary sources were:

1. ECDC: European Centre for Disease Control. Data downloaded from <https://www.ecdc.europa.eu/en/publications-data/weekly-subnational-14-day-notification-rate-covid-19> & https://www.ecdc.europa.eu/en/publications-data/data-national-14-day-notification-rate-covid-19
2. OWID: Our World in Data-COVID-19: Downloaded from https://github.com/owid/covid-19-data/tree/master/public/data

Other sources stated below.

*Best estimate used first the data reported in the paper and if not available, the mean of 14-day incidence across the weeks of the study

| **Authors** | **Country** | **Dates** | **Week number** | **Subnational region** | **Source** | **Data from paper** | **14-day contemporary incidence best estimate*** | **14-day incidence contemporary to study** | **14-day incidence for month prior to study** | **14-day incidence 2 months prior to study** |
| --- | --- | --- | --- | --- | --- | --- | --- | --- | --- | --- |
| Villani et al A | Italy | 21 Sep-12 Oct 2020 | 39-41 | Lazio | ECDC |  | 61.81119007 | 61.81119007 | 38.07529927 | 10.5286933 |
| Villani et al B | Italy | 19 Oct-13 Nov 2020 | 43-46 | Lazio | ECDC |  | 463.162604 | 463.162604 | 85.9669545 | 38.07529927 |
| Villani et al C | Italy | 16 Nov-4 Dec | 47-49 | Lazio | ECDC |  | 560.3662456 | 560.3662456 | 463.162604 | 85.9669545 |
| Brandal et al. | Norway | 28 Aug-11 Nov 2020 | 36-46 | Oslo and Viken counties | Paper & ECDC | 19.3 to 94.9 cases per 100,000 for Week 36-46 | 94.9 | 97.66718839 | 24.36612527 | 6.153895675 |
| Schoeps et al. | Germany | 17 Aug-16 Dec 2020 | 34-51 | Rhineland-Palatinate | ECDC | 120 to 170 per 100,000 | 145 | 143.6187206 | 10.38739804 | 4.812033895 |
| Hu et al. | China (Hunan) | 23 Jan-2 April 2020 | 4-14 | - | OWID |  | 0.518290909 | 0.518290909 |  |  |
| Kriemler et al | Switzerland | 1-11 Dec 2020 | 49-50 |  | [ECDC & https://www.covid19.admin.ch/](https://www.covid19.admin.ch/) | 300 per 100’000 new cases per day | 639.7857143 | 639.7857143 | 438.4249979 | 438.4249979 |
| Ulyte et al.(A) | Switzerland | 16 Jun-9 July | 25-28 |  | [ECDC& https://www.covid19.admin.ch/](https://www.covid19.admin.ch/) | Seroprevalence of adults in same distict in June 2020=3.1%(1.4, 5.4) | 10.85 | 10.85 | 3.456877286 | 22.79796045 |
| Ulyte et al.(B) | Switzerland | T2 26 Oct-19 Nov | 44-47 |  | [ECDC& https://www.covid19.admin.ch/](https://www.covid19.admin.ch/) | 590 daily cases per million Nov 2020 | 826 | 986.688 | 222.8378627 | 56.14375404 |
| Hommes et al. | Germany | 11-19 Jun 2020 | 25 | Berlin | ECDC | 3-14/100,000 | 14 | 24.33580025 | 11.80000169 | 17.65912493 |
| Theuring et al. | Germany | 2-16 Nov 2020 | 45-46 | Berlin | ECDC | 185-210/100,000 | 197.5 | 383.2275376 | 208.727859 | 51.17194728 |
| Armann et al1 | Germany | Time 1 25 May-30 June 2020 | 22-27 | Saxony | Paper & ECDC | T1: 139/ 100,000; | 139 | 2.193859763 | 9.810973605 | 34.88482605 |
| Armann et al.2 | Germany | Time 2: 15 Sep-13 Oct 2020 | 38-42 | Saxony | Paper & ECDC | T2: 245/100,000 | 245 | 34.33226808 | 7.588462688 | 2.707534017 |
| Thielecke et al. | Germany | 28 Sep-2 Oct 2020 | 40 | Berlin | ECDC | 7 day incidence 38/100,000 | 76 | 78.02172018 | 38.66340046 | 25.40543089 |
| Hoch et al.1 | Germany | Time 1: 15 Jun-26 July | 25-29 | Bayern | ECDC | - | 6.387937526 | 6.387937526 | 9.442093963 | 31.00252599 |
| Hoch et al. 2 | Germany | Time 2: 7 Sep-1 Nov 2020 | 37-44 | Bayern | Paper & ECDC | T2: 150/100,000 | 150 | 86.49601893 | 27.32054745 | 9.86495958 |
| Willeit et al.A | Austria | Time 1: 28 Sep-22 Oct 2020 | 40-43 | - | Paper & ECDC | T1: 75/100,000; | 75 | 185.5677029 | 83.04063424 | 29.94585816 |
| Willeit et al.B | Austria | Time 2: 10-16 Nov 2020 | 46 | - | Paper & ECDC | T2: 419/100,000 | 419 | 1037.898391 | 446.6235722 | 118.1656485 |
| Varma et al.1 | USA | Period 1 9 Oct-20 Nov | 41-47 | NYC | ECDC & https://github.com/nychealth/coronavirus-data/blob/a95893030e94cffa0e7349f083a1717a67546822/trends/hosprate-by-modzcta.csv | Period 1: 529/100,000 | 529 | 529 | 170.641158 | 193.4097071 |
| Varma et al. 2 | USA | Period 2: 6-18 Dec 2020 | 50-51 | NYC | ECDC & https://github.com/nychealth/coronavirus-data/blob/a95893030e94cffa0e7349f083a1717a67546822/trends/hosprate-by-modzcta.csv | Period 2: 510/100,000 | 510 | 510 | 676.830388 | 302.008461 |
| Reukers et al. | Netherlands | Mar-May 2020 | 10-22 | Utrecht | ECDC |  | 142.2635009 | 39.64430793 | 39.64430793 | 39.64430793 |
| Dattner et al. | Israel | 17 Mar-3 May 2020 | 12-18 | - | OWID |  | 26.27382857 | 26.27382857 | 0.511325 | 0.511325 |
| Larosa et al. | Italy | 1 Sep-15 Oct 2020 | 36-42 | Reggio Emilia | ECDC |  | 54.90765047 | 54.90765047 | 20.66477171 | 12.47166574 |
| Pray et al. | USA | July-Aug 2020 |  |  |  |  |  |  |  |  |
| ONS SIS1 | UK | Round 1: 3-19 Nov 2020; | 45-47 | - | OWID | England Round 1: 230/100,000 | 230 | 234.584 | 196.07685 | 57.493825 |
| ONS SIS2 | UK | Round 2: 2-10 Dec 2020 | 49-50 | - | OWID |  | 208.6125 | 208.6125 | 214.791025 | 196.07685 |
| Kim et al. | South Korea | 20 Jan-6-Apr 2020 | 4-14 | - | OWID & ECDC |  | 3.13215 | 3.13215 | 0.003250816 | 0.003250816 |
| Maltezou et al. | Greece | 26 Feb-3 May 2020 | 9-18 | - | OWID & ECDC |  | 2.5194 | 2.5194 | 0.0001 | 0.0001 |
| Laxminarayan et al. | India | 5 Mar-June 2020 | 11-25 | - | OWID |  | 2.0543 | 2.0543 | 0.00065 | 0.00065 |
| Lyngse et al. | Denmark | 25 Aug 2020-10 Feb 2021 | 35(2020)-7(2021) |  | OWID & ECDC |  | 117.4468492 | 117.4468492 | 22.03850646 | 6.63344876 |
| Ladhani et al. 1 | UK | RT-PCR June-July. Serology round 1 June, | 23-27 |  | OWID |  | 8.65924 | 8.65924 | 24.979775 | 47.24325 |
| Ladhani et al.2 | UK | Serology round 2 July 2020 | 27-31 |  | OWID |  | 6.51594 | 6.51594 | 9.815375 | 24.979775 |
| Ladhani et al.3 | UK | Serology round 3 Nov-Dec 2020. | 45-52 |  | OWID |  | 231.4866375 | 231.4866375 | 196.07685 | 57.493825 |
| Fontanet et al. | France | 28-30 April 2020 | 18 | Ile de France | ECDC |  | 60.50983665 | 60.50983665 | 60.50983665 | 60.50983665 |
| Jordan et al. | Spain | 29- Jun - 31 July 2020 | 27-31 | Cataluna | ECDC |  | 97.12835851 | 97.12835851 | 23.93056353 | 48.40671125 |
| Yoon et al. | South Korea | 20 May-31 July 2020 | 21-31 | - | OWID | May 2020: 21/100,000 | 21 | 21 | 0.197125 | 0.56325 |
| Macartney et al.T3 | Australia | Term3 ( 4 July-25 Sep ), | 28-39 | - | OWID |  | 6.033008333 | 6.033008333 | 1.292175 | 0.310775 |
| Macartney et al.T4 | Australia | Term 4 (26 Sep-18 Dec). | 40-51 | - | OWID |  | 0.377133333 | 0.377133333 | 1.272475 | 7.282325 |
| Blaisdell et al. | USA | June-August 2020 | 23-35 | Maine | https://covid.cdc.gov/covid-data-tracker & https://www.maine.gov/dhhs/mecdc/infectious-disease/epi/airborne/coronavirus/data.shtml |  | 23.77225881 | 23.77225881 |  |  |
| Park et al. | South Korea | 20 Jan-27 Mar 2020 | 4-13 | National | OWID |  | 1.86903 | 1.86903 | 1.86903 | 1.86903 |
| Li et al. | Wuhan, China | 2 Dec-18 Apr 2020 | 1-16 |  | OWID | 0.444992308 | 0.444992308 | 0.4523 | 0.444992308 | 0.444992308 |
| House et al. 3 | UK | 15 Nov -31 Dec 2020 | 47-52 | National | ECDC |  | 442.8811932 | 442.8811932 | 453.7204255 | 206.9369431 |
| House et al. 4 | UK | 1 Jan-15 Feb 2021 | 1-7 | National | ECDC |  | 665.1870994 | 665.1870994 | 603.8581065 | 409.0670885 |
| Lubke | Germany | 10 Jun - 7 July 2020 | 24-28 | Nordrhein-Westfalen | ECDC | Prevalence: 0.81 infections per 1,000 inhabitants | 13.9397626 | 13.9397626 | 10.75375402 | 32.79477085 |
| Hoehl | Germany | 18 Jun-10 Sept | 25-37 | Hessen | ECDC | Peaks up to 66/100,000 | 33 | 15.67923876 | 8.917666442 | 18.0858068 |
| Telle et al. | Norway | 1 March 2020-1 Jan 2021 | 10-52 | - | ECDC |  | 38.91277668 | 38.91277668 | 21.22744328 | 28.94693698 |
| Espenhain2 | Denmark | R2 August 2020 | 31-35 |  | ECDC |  | 21.35068867 | 21.35068867 | 6.63344876 | 8.745847976 |
| Espenhain3 | Denmark | Oct-20 | 40-44 |  | ECDC |  | 134.5443735 | 134.5443735 | 67.22324093 | 24.04356832 |
| Espenhain4 | Denmark | Dec-20 | 49-52 |  | ECDC |  | 591.550781 | 591.550781 | 266.9780652 | 134.5495257 |
| Doron1 | USA | 16-Sep-20 | 38 |  |  | 14d incidence per 100,000: Baseline: 28 | 28 | 70.53103134 | 4.270167491 | 4.270167491 |
| Doron2 | USA | 1 Oct to 20 Nov 2020 | 40-47 |  | https://covid.cdc.gov/covid-data-tracker/#trends_dailytrendscases | 221 | 221 | 256.136802 | 77.17576144 | 75.31496733 |
| Doron3 | USA | 7-31 Dec 2020 | 50-52 |  |  | 352 | 352 | 920.7146613 | 430.1940234 | 161.5993986 |
| SIS4A | UK | 15-30 March 2021 | 12-14 2021 |  | ECDC |  | 87.03612387 | 87.03612387 | 147.4562181 | 407.789904 |

#### Appendix Table 6. Data on attendance in school studies

| **Authors** | **School mitigations in place reported in paper** | **Face to face attendance %** |
| --- | --- | --- |
| Villani et al | administrative policies, infrastructural adjustments, sanitation of environments, appropriate use of individual protection devices, symptoms screenings by parents and teachers | 100 |
| Brandal et al. | strengthened hygiene, physical distancing; masks not used | 100 |
| Schoeps et al. | Secondary: physical distancing (> 1·5 meters); cross- or pulse-ventilation of class-rooms before and after class, and then every 20 minutes during class,; face masks in school-buildings & in the class-room from Nov; increased surface cleaning. Primary & day-care- similar without distancing and masks | 100 |
| Kriemler et al | masks for teachers and children >12-years-old in communal areas (not classrooms), physical distancing, staggered school breaks, classes in bubbles and no gatherings of multiple classes, no parents on school grounds | 100 |
| Ulyte et al. | R1: not stated. R2: masks for teachers and children >12-years-old in communal areas (not classrooms) in Sept 2020; physical distancing, staggered school breaks, classes in bubbles and no gatherings of multiple classes, no parents on school grounds. R3: As previously plus masks for all students | 100 all R |
| Hommes et al. | improved ventilation; physical distancing; facemask use in communal areas and also in class in class some schools; class cohorting; sports cancelled; Limited pupil numbers and reduced schedules: on average 15% of learning was online in primary schools and 50% at secondary; | 60 |
| Theuring et al. | enhanced ventilation; class cohorting; facemasks in communal areas in all; masks in class in 2/3 of schools; hand hygiene. Some online teaching in 11/22 schools (not specified) | 90 |
| Kirsten et al. | Not stated. Data copied from other German studies on same dates. | R1=90, R2=100 |
| Thielecke et al. | physical distancing; staff facemask; cohorting of children; enhanced ventilation | 100 |
| Hoch et al. | Time 1: physical distancing; enhanced hygiene; cancellation common activities. Time 2: physical distancing; enhanced hygiene; staff facemask; reduced parental visiting; cancellation common activities | 100 both times |
| Willeit et al. | Round 1 & 2 Physical distancing; cohorting of classes in bubbles; increased ventilation; sports only with 2m distancing. Masks: R1 - largely no masks; R2 masks in communal areas | 100 both times |
| Varma et al. | Both rounds for in-school learning: reduced class sizes and cohorting of classes; Temperature checks; symptom screening; enhanced ventilation; masks at all times; physical distancing; exclusion of visitors; improved ventilation | Round 1=20%; Round 2=100% |
| Larosa et al. | physical distancing; mandatory masks except at desks in secondary (no masks primary); cohorting of classes and staggered start and break times; some limitation in size of classes; no sports or music | unclear but not 100%; estimate 90% |
| SIS | R1 & 2: increased hand sanitising; increased cleaning; masks in communal areas in secondary (not classrooms); class cohorting in bubbles; R4 & R5: also included masks in classrooms | 100 |
| Ladhani sKids | Schools FTF: Strict physical distancing and infection control measures implemented, including smaller class sizes, cohorting of staff and students, enhanced cleaning | 100 |
| Fontanet et al. | No mitigations in place | 100 |
| Jordan et al. | bubble groups, hand washing of 8-14 children, facemasks and conducting activities mostly outdoors | 100 |
| Yoon et al. | Hybrid learning, restrictions on numbers in classrooms, physical distancing, masks at all times except playground, music lessons suspended | 67 |
| Macartney et al. | Term 3: hand hygiene, no parents or visitors, enhanced cleaning, cohorting of year groups, physical distancing, singing and group activities restricted. Term 4: minimal restrictions - all activities allowed - distancing only required for musical activities | 100 |
| Blaisdell et al. | Prearrival quarantine, pre- and post-arrival testing and symptom screening, cohorting of residential groups for indoor activities, use of face coverings, physical distancing, enhanced hygiene measures, cleaning and disinfecting, and maximal outdoor programming. Face coverings and physical distancing if mixing outside cohort for 1st 14 days. All attendees quarantined with families for 14 days before arrival and for 14 days with their residential cohort post-arrival. Temperature and symptom screening daily. |  |
| Lubke et al. | Not stated | 100 |
| Hoehl et al./ Schenk | R1: Limited parent access; hygiene and cleaning measures; R2 & R3 not stated | 100 all rounds |
| Doron et al. | Hybrid learning began 1 October: all attended 2 days FTF+2.5 days remote, except K-2 and students with high learning needs who attended FTF 4 days per week.. In person: mandatory masks in all areas, frequent hygiene, physical distancing, upgraded ventilation, daily symptom screening | Round 1: 0 Round 2: 50 Round 3: 50 |
| Ladhani sKidsPLUS | Schools FTF: Strict physical distancing and infection control measures implemented, including smaller class sizes, cohorting of staff and students, enhanced cleaning; masks in class | 100 |
| Lachassine | strict sanitary measures were introduced and enforced in the daycare centres; children were hosted in small, unchanging groups, looked after by a single constant worker for a week; enhanced cleaning; staff wearing masks and social distancing; parents and visitors excluded; symptomatic screening of children | 100 |

#### Appendix Table 7. Studies excluded due to high risk of bias

##### A. Contact tracing studies (Household (HH) and School)

| Authors | Reference/DOI | Setting | Country | Description | Reason for exclusion |
| --- | --- | --- | --- | --- | --- |
| 1. Lopez-Bernal et al. | <https://doi.org/10.1101/2020.08.19.20177188> | HH | UK | Prospective CTS undertaken in England early in the pandemic in February and March 2020. | Symptomatic testing of contacts |
| 2. Chu et al. | <https://doi.org/10.1101/2020.10.10.20210492> | HH of attendees of School residential camp | USA | CTS amongst HH contacts of children from an overnight camp. 72% of contacts tested. | Symptomatic testing of contacts |
| 3. Wang et al. | https://doi.org/10.1101/2020.10.22.20217661 | Community | USA | used state-wide contact-tracing data from Georgia from 1 February to 13 July, to identify age-based transmission amongst 4080 transmission pairs within a 14-day interval. |  |
| 4. Cordery et al. | https://doi.org/10.1101/2021.03.08.21252839 | HH & schools | UK | CTS in HH contacts of 5 index cases from London schools in October-December 2020 | Participation of school contacts was 30%, with bubble contacts at 15%. |
| 5. Grijalva et al. | https://www.cdc.gov/mmwr/volumes/69/wr/mm6944e1.htm | HH | USA | Used routine surveillance data from two US cities to undertake a detailed study of transmission in the households of 101 index cases, of whom 14% were under 18y. | All contacts were tested however participation rates for HH and contacts not stated. |
| 6. Laws et al. | https://doi.org/10.1542/peds.2020-027268 | HH | USA | CTS in in a convenience sample of 33 US HH in 2 cities with resident children. All contacts were tested. | One index case < 18y i.e. similar to case report for transmission from children. Proportion of contacts who participated not stated. |
| 7. Van de Hoek et al. | https://www.ntvg.nl/artikelen/de-rol-van-kinderen-de-transmissie-van-sars-cov-2 | HH | Netherlands | Used Dutch national surveillance data to identify transmission amongst 732 PCR-positive ‘pairs’ that lived in the same household | Symptomatic testing of contacts |
| 8. Posfe-Barbe et al. | https://doi.org/10.1542/peds.2020-1576 | HH | Switzerland | CTS in all HH with positive child <16y in Geneva. | Predominantly symptomatic testing of contacts (58% tested) |
| 9. Somekh et al. | <https://journals.lww.com/pidj/Fulltext/2020/08000/The_Role_of_Children_in_the_Dynamics_of_Intra.30.aspx> | HH | Israel | CTS of 13 HH clusters in Bnei Brak. All contacts were tested. | Potential duplicate of data contained in Dattner et al. |
| 10. Yoon et al. | <https://doi.org/10.3201/eid2702.203189> | School | South Korea | CTS of contacts of a child who attended an early-years setting in South Korea while pre-symptomatic. | Single index case. Note tested all contacts |
| 11. Ehrhardt et al. | <https://www.eurosurveillance.org/content/10.2807/1560-7917.ES.2020.25.36.2001587> | School | Germany | CTS of cases attending school whilst symptomatic in state of Baden-Wurttemberg. | Large number of contacts tested however testing strategy, % of contacts recruited and tested not stated. |
| 12. Macartney et al. | <https://doi.org/10.1016/S2352-4642(20)30251-0> and <https://www.ncirs.org.au/reports> | School | Australia | School-based surveillance and contact tracing of positive cases attending school while symptomatic in State of New South Wales. Data were collected for Terms 1-4 of 2020. Term 3 & 4 data are included in the review but Terms 1 and 2 excluded. | Term 1 data. Symptom-based testing of contacts - 44% of contacts were tested. Enhanced surveillance cohort (7 schools) data also not eligible as 67% of contacts were tested by PCR, serology or both.  Term 2 data: predominantly symptom-based testing: 61% of contacts had PCR. |
| 13. Falk et al. | https://www.cdc.gov/mmwr/volumes/70/wr/mm7004e3.htm | School | USA | School-based contact-tracing of all identified symptomatic infections in 17 schools in Wisconcin, USA in Sept-Nov 2020. | Symptomatic testing of contacts |
| 14. Zimmerman et al. | <https://doi.org/>10.1542/peds.2020-048090 | School | USA | Undertook a CTS in 11 North Carolina school districts in the first 9 weeks of school resumption in August 2020. | Symptomatic or non-systematic testing of contacts |
| 15. Heudorf et al. | https://pubmed.ncbi.nlm.nih.gov/33214989/ | Community and schools | Germany | Used routine public health data from Frankfurt, Germany, during March to July 2020 to examine transmission from child index cases | Symptomatic case and contact identification |
| 16. Fong et al. | <https://www.eurosurveillance.org/content/10.2807/1560-7917.ES.2020.25.37.2001671#html_fulltext> | Schools | Hong Kong | Reported national surveillance data on schools in Hong Kong after schools reopened in late May (secondary) and early June (primary). | Lack of detail on testing of contacts; numbers of contacts tested not stated |
| 17. Heavy et al. | https://doi.org/10.2807/1560-7917.ES.2020.25.21.2000903 | Schools | Ireland | National contact-tracing study in the Republic of Ireland before schools closed on 12 March 2020 | Symptomatic testing of contacts |
| 18. Yung et al. | https://doi.org/10.1093/cid/ciaa79 | Schools | Singapore | National CTS in schools in Singapore using national surveillance registry data, in February and March 2020 before schools closed. | Symptomatic testing of contacts |
| 19. Dub et al. | <https://www.medrxiv.org/content/10.1101/2020.07.20.20156018v1> | Schools | Finland | Studied transmission from two child cases identified in schools in Helsinki in March 2020 | Case series; only 67% of child contacts tested |
| 20. Lopez et al. | <https://www.cdc.gov/mmwr/volumes/69/wr/mm6937e3.htm> | Child-care facilities | USA | Surveillance and contact-tracing in child-care facilities in Salt Lake City 1 April -10 July 2020 | Symptomatic testing only |
| 21. Kriger et al. | <https://doi.org/10.1016/j.cmi.2020.11.030> | Schools | Israel | CTS of children exposed to an infected teacher in an alternative school | Single index case |
| 22. Gold et al. | https://www.cdc.gov/mmwr/volumes/70/wr/mm7008e4.htm?s_cid=mm7008e4_w | Schools | USA | CTS followed identified cases in a Georgia school district during December 1, 2020–January 22, 2021 | 60% of contacts tested |
| 23. Lewis et al. | <https://doi.org/10.1093/cid/ciaa1166> | HH | USA | HH CTS in Utah and Wisconsin During March-April 2020 | Single index case <18y |
| 24. Maltezou et al. | <https://pubmed.ncbi.nlm.nih.gov/32767703/> | HH | Greece | Contact-tracing of 23 family clusters in Greece during Feb-May 2020. | Lack of clarity on mapping of contacts to index cases |

#### B. School surveillance studies

| Authors | Reference/DOI | Setting | Country | Description | Reason for exclusion |
| --- | --- | --- | --- | --- | --- |
| 25. Gandini et al. | https://doi.org/10.1101/2020.12.16.20248134 | School | Italy | Used Italian national public health and educational surveillance data to estimate infection incidence and secondary infection rates in school-children | Symptomatic testing only |
| 26. Ismail et al. | https://www.ncbi.nlm.nih.gov/pmc/articles/PMC7833602/ | School | England | National surveillance data involving educational settings after schools and early-years settings in England partially reopened with mitigations in place from 1 to 30 June 2020. | Predominantly symptomatic testing; some wider testing in schools during outbreaks |
| 27. Wada et al. | <https://bmjpaedsopen.bmj.com/content/4/1/e000854> | School | Japan | Used Japanese national surveillance data from primary and junior secondary schools to study cases and transmission in schools | Symptomatic testing only |
| 28. Otte im Kampe et al. | <https://www.eurosurveillance.org/content/10.2807/1560-7917.ES.2020.25.38.2001645#html_fulltext> | School | German | Used German national surveillance data to examine cases and outbreaks in schools | Symptomatic testing predominantly |
| 29. Cornelissen et al. | <https://covid-19.sciensano.be/sites/default/files/Covid19/COVID-19_THEMATIC%20REPORT_COVID-19%20INFECTION%20IN%20CHILDREN_FR.pdf> | School and community | Belgium | Used Belgian national surveillance data on infections in school-children undertaken by the national health institute Sciensano | Symptomatic testing only |
| 30. Link -Gelles et al. | <https://www.cdc.gov/mmwr/volumes/69/wr/mm6934e2.htm> | Early years settings | USA | Data from public health surveillance of all potential cases from 66 child-care facilities in Rhode Is, USA. | Symptomatic testing only |
| 31. Russell et al. | https://www.mcri.edu.au/sites/default/files/media/covid_in_schools_report_final_10112020.pdf | Schools and EYS | Australia | Public health surveillance of school and early years settings related cases and contact-tracing | Symptomatic testing predominantly |
| 32. Thompson et al. | https://doi.org/10.1101/2021.02.04.21251087 | Schools | Wales | Used national linkage of school and PCR-testing databases in Wales to examine temporal associations of symptomatically-identified infections with subsequent infections by class and school. | Symptomatic testing predominantly |
| 33. Frank et al. | <https://doi.org/10.1101/2021.04.03.21254873> | Early years setting | USA | Weekly testing of children in an early years setting. | Focuses on validation of salivary sampling. Insufficient data to include |
| 34. Cooper et al. | <https://doi.org/10.1101/2021.03.20.21254035> | Schools | USA | Longitudinal testing of subset of learners in 4 schools | Non-random small sample of school children in each school. Unclear if representative. |
| 35. Gillespie et al. | <https://doi.org/10.1111/josh.13008> | Schools | USA | Longitudinal testing of students and staff in 2 schools | Not population based |
| 36. Llupià et al. | <https://doi.org/10.1371/journal.pone.0251593> | Schools | Catalonia, Spain | Analysis of infections within schools in Catalonia, | Unclear testing policy. Likely symptomatic testing predominantly |
| 37. Mossong et al. | <https://doi.org/10.1186/s12879-021-06089-5> | Schools | Luxembourg | Public health surveillance of school infections in early summer 2020 | Number of contacts not stated; proportion of contacts tested not stated although all offered testing |
| 38. Nelson et al. | <https://doi.org/10.1001/jama.2021.2392> | Schools | Florida, USA | Contact-tracing study in schools | Proportion of contacts tested unclear |
| 39. White et al. | <https://doi.org/10.1016/j.puhe.2021.04.001> | Schools | Ireland | Public health surveillance of school infections | Predominantly symptomatic testing of contacts |
| 40. Lanier et al. | <https://www.ncbi.nlm.nih.gov/pmc/articles/PMC8158889/pdf/mm7021e2.pdf> | Schools | Utah, USA | Longitudinal rapid antigen testing in schools in Utah | Did not use RT-PCR for antigen identification – used rapid antigen tests |
| 41. Haag et al. | <https://doi.org/10.1101/2021.04.16.21255616> | Child care settings | Germany | Infection prevalence in childcare settings in Saxony, German | Did not use nasopharyngeal or oral swab/salivaRT-PCR for antigen identification – stool samples used. |
| 42. Cooper et al. | <https://doi.org/10.1101/2021.03.20.21254035> | Schools | California | Prospective study of four purposively-chosen schools in Orange County, California. | Not population-based |
| 43. Bark et al. | <https://doi.org/10.1101/2021.05.15.21257271> | Schools | Vancouver, Canada | Contact-tracing study in K-12 schools in Vancouver region | Symptomatic testing of contacts |
| 44. Loenenbach et al. | <https://doi.org/10.1101/2021.05.12.21256608> | Kindergarten | Germany | Contact-tracing study of outbreak in 3 kindergartens with testing of households | Households were not tested systematically. Proportion of contacts tested not stated. |
| 45. Jurkutat et al. | <https://home.uni-leipzig.de/lifechild/schulerhebung-corona/> | Schools | Germany | Surveillance of infection with SARS-CoV-2 among teachers, students in Saxony | Lack of detail on response rates and detail on numerators and denominators |

##### Appendix Figure 1. Forest plot of secondary attack rate (SAR) from child index cases to child compared with adult contacts in school contact-tracing studies

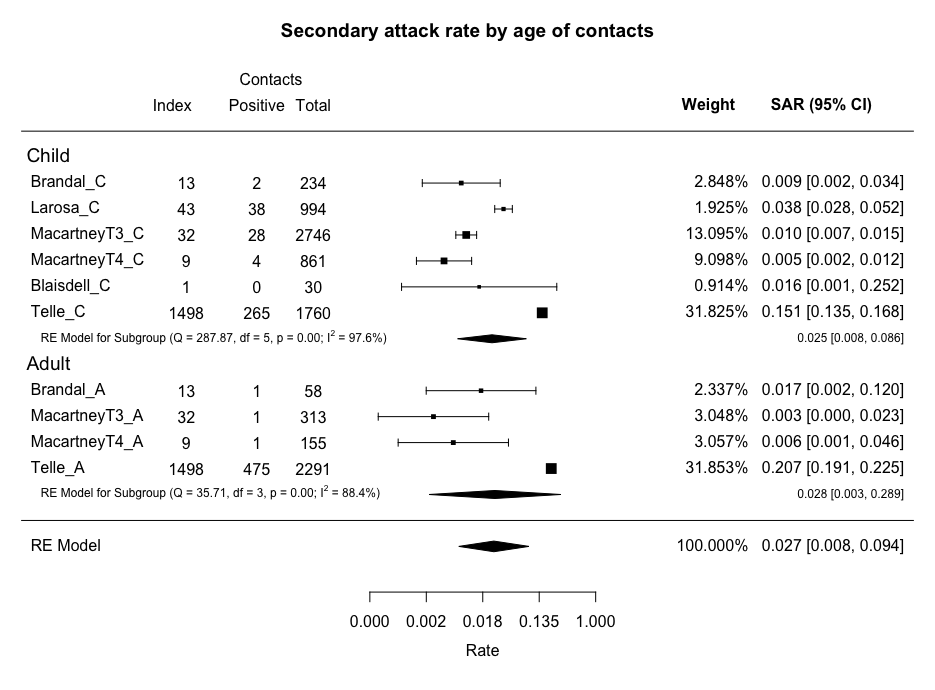
